# A robust, highly multiplexed mass spectrometry assay to identify SARS-CoV-2 variants

**DOI:** 10.1101/2022.05.28.22275691

**Authors:** Matthew M. Hernandez, Radhika Banu, Paras Shrestha, Ana S. Gonzalez-Reiche, Adriana van de Guchte, Keith Farrugia, Robert Sebra, Mount Sinai PSP Study Group, Melissa R. Gitman, Michael D. Nowak, Carlos Cordon-Cardo, Viviana Simon, Harm van Bakel, Emilia Mia Sordillo, Nicolas Luna, Angie Ramirez, Sergio Andres Castañeda, Luz Helena Patiño, Nathalia Ballesteros, Marina Muñoz, Juan David Ramírez, Alberto E. Paniz-Mondolfi

## Abstract

Severe acute respiratory syndrome coronavirus 2 (SARS-CoV-2) variants are characterized by differences in transmissibility and response to therapeutics. Therefore, discriminating among them is vital for surveillance, infection prevention, and patient care. While whole viral genome sequencing (WGS) is the “gold standard” for variant identification, molecular variant panels have become increasingly available. Most, however, are based on limited targets and have not undergone comprehensive evaluation. We assessed the diagnostic performance of the highly multiplexed Agena MassARRAY^®^ SARS-CoV-2 Variant Panel v3 to identify variants in a diverse set of 391 SARS-CoV-2 clinical RNA specimens collected across our health systems in New York City, USA as well as in Bogotá, Colombia (September 2, 2020 – March 2, 2022). We demonstrate almost perfect levels of interrater agreement between this assay and WGS for 9 of 11 variant calls (κ ≥ 0.856) and 25 of 30 targets (κ ≥ 0.820) tested on the panel. The assay had a high diagnostic sensitivity (≥93.67%) for contemporary variants (e.g., Iota, Alpha, Delta, Omicron [BA.1 sublineage]) and a high diagnostic specificity for all 11 variants (≥96.15%) and all 30 targets (≥94.34%) tested. Moreover, we highlight distinct target patterns that can be utilized to identify variants not yet defined on the panel including the Omicron BA.2 and other sublineages. These findings exemplify the power of highly multiplexed diagnostic panels to accurately call variants and the potential for target result signatures to elucidate new ones.

**Importance:** The continued circulation of SARS-CoV-2 amidst limited surveillance efforts and inconsistent vaccination of populations has resulted in emergence of variants that uniquely impact public health systems. Thus, in conjunction with functional and clinical studies, continuous detection and identification are quintessential to inform diagnostic and public health measures. Furthermore, until WGS becomes more accessible in the clinical microbiology laboratory, the ideal assay for identifying variants must be robust, provide high resolution, and be adaptable to the evolving nature of viruses like SARS-CoV-2. Here, we highlight the diagnostic capabilities of a highly multiplexed commercial assay to identify diverse SARS-CoV-2 lineages that circulated at over September 2, 2020 – March 2, 2022 among patients seeking care at our health systems. This assay demonstrates variant-specific signatures of nucleotide/amino acid polymorphisms and underscores its utility for detection of contemporary and emerging SARS-CoV-2 variants of concern.

## Introduction

Since the onset of the coronavirus disease 2019 (COVID-19) pandemic, suboptimal surveillance and diagnostic efforts have not been able to prevent the rapid, unchecked spread of severe acute respiratory syndrome coronavirus 2 (SARS-CoV-2) (1–4). In conjunction with various factors (e.g., variable healthcare access, limitations to effective infection prevention efforts), continued spread has led to the emergence of viral variants characterized by increased genomic diversity including the most recent Omicron (B.1.1.529) variant and its sublineages (5–8). This poses a unique challenge to healthcare systems and diagnostic laboratories, alike, as genomic variation has the potential to impact viral fitness (5, 9), disease pathogenesis (10–12), response to therapeutics (e.g., antibodies) (5, 13, 14), and molecular target detection (15–17).

Ideally, SARS-CoV-2 diagnostic assays should be scalable to test increased clinical specimens and should be robust enough to accommodate genomic variation in viruses over time. Although improved technologies have made high-throughput platforms more available, most are limited in the level of multiplexing and, thus, risk target dropout and failing to capture infected individuals. Indeed, current nucleic acid amplification tests (e.g., reverse-transcription polymerase chain reaction (RT-PCR)) largely utilize 1-3 targets to detect (e.g., presence/absence) SARS-CoV-2 nucleic acids. Moreover, as new variants have emerged, diagnostic panels are based on targets designed for detection of nucleotide changes that yield specific amino acid substitutions and call variants based on distinct target result combinations (18–20). However, most of these assays distinguish viral variants through result patterns of 3-9 molecular targets across multiple reaction wells (20–28), which are constrained to distinguishing current circulating variants but may not be sufficient to distinguish nascent, increasingly divergent variants.

Whole viral genome sequencing (WGS) has, therefore, largely served as the “gold standard” for pathogen genomic surveillance. Still, this methodology is not realistic for most diagnostic laboratories as it requires staff with bioinformatic expertise, bioinformatics infrastructure, is relatively expensive and restricted in lower income countries (LICs) and lower middle income countries (LMCs) (29, 30). Therefore, there is great potential for highly multiplexed assays that target an expansive repertoire of polymorphisms. Currently, these platforms are rare in number (31, 32) and most have not yet been evaluated for their diagnostic capabilities in the clinical setting.

Here, we recovered 391 SARS-CoV-2 viral RNA from clinical specimens collected from infected individuals who presented for testing at the Mount Sinai Health System (MSHS) in New York City (NYC) and at the Universidad del Rosario in Bogotá, Colombia from September 2, 2020 – March 2, 2022. These specimens had previously undergone WGS for epidemiologic surveillance, and we used this data as a benchmark to evaluate the diagnostic performance of the Agena MassARRAY^®^ SARS-CoV-2 Variant Panel v3 (research use only, RUO). We tested this highly diverse set of viral variants to interrogate the level of agreement and diagnostic sensitivity and specificity across 12 distinct variants on the panel and 30 distinct polymorphic targets in the *Spike (S)* gene region. We demonstrate a high level of assay agreement and high levels of diagnostic sensitivity and specificity across most variant and individual targets tested. Furthermore, we highlight the utility of the variant panel to elucidate undefined or emergent variants based on unique target result signatures.

## Materials and Methods

### Ethics statement

For specimens obtained through routine testing at MSHS, the Mount Sinai Pathogen Surveillance Program was reviewed and approved by the Human Research Protection Program at the Icahn School of Medicine at Mount Sinai (ISMMS) (HS#13-00981). For specimens from Colombia, the study was reviewed and approved by the ethics Committee from Universidad del Rosario in Bogotá, Colombia (Act number DVO005 1550-CV1499). This study was performed following the Declaration of Helsinki and its later amendments, and all patient data was anonymized to minimize risk to participants.

### SARS-CoV-2 specimen collection and testing

Residual viral RNA from a total of 391 specimens that were previously collected from September 2, 2020 – March 2, 2022 for routine diagnostic testing were utilized for this study.

Specifically, 349 upper respiratory tract (e.g., nasopharyngeal, anterior nares) and saliva (September 2, 2020 – March 2, 2022) specimens were originally collected for SARS-CoV-2 diagnostic testing in the Molecular Microbiology Laboratory of the MSHS Clinical Laboratory, which is certified under Clinical Laboratory Improvement Amendments of 1988, 42 U.S.C. §263a and meets requirements to perform high-complexity tests were eligible for inclusion in this study. Viral RNA was extracted from 300μL of each specimen using the Viral DNA/RNA 300 Kit H96 (PerkinElmer, CMGL1033LS) on the automated chemagic(tm) 360 instrument (PerkinElmer, 2024L0020) per manufacturer’s protocol as previously described (33, 34). After routine testing and extraction, viral RNA was stored at -80°C prior to recovery for testing in this study.

Forty-two nasopharyngeal specimens were collected from patients from the Valle del Cauca department for SARS-CoV-2 testing at Universidad del Rosario from March 29, 2021 – July 28, 2021. Details of processing and SARS-CoV-2 testing of upper respiratory specimens have been described previously (35). After diagnostic testing, residual viral RNA was also stored at -80°C prior to recovery for testing in this study.

### SARS-CoV-2 sequencing, assembly, and phylogenetics

As part of the ongoing Mount Sinai Pathogen Surveillance Program, SARS-CoV-2 viral RNA from MSHS underwent RT-PCR and next-generation sequencing followed by genome assembly and lineage assignment using a phylogenetic-based nomenclature as described by Rambaut et al. (36) using the Pangolin v4.0.6 tool and PANGO-v1.2.81 nomenclature scheme (https://github.com/cov-lineages/pangolin) as previously described (4, 37).

Sequence libraries were prepared from RNA from Colombian specimens using the ARTIC Network protocol (https://artic.network/ncov-2019 accessed on 1 February 2021) as previously described (38). Briefly, long-read Oxford Nanopore MinION sequencing was conducted by the MinKNOW application (v1.5.5). Raw Fast5 files were base called and demultiplexed using Guppy. Reads were filtered to remove possible chimeric reads, and genome assemblies were obtained following the MinION pipeline described in the ARTIC bioinformatics pipeline (https://artic.network/ncov-2019/ncov2019-bioinformatics-sop.html accessed on 1 February 2021).

Of 391 specimens, 381 single variant consensus genome sequences were identified, and the remaining 10 yielded mixed assemblies and, thus, a putative (inconclusive) consensus genome sequence was generated. All FASTA consensus genome sequences underwent mutation calling and phylogenetic lineage assignment by the Nextclade Web Interface (https://clades.nextstrain.org/, last accessed 4/18/22) and the Pangolin COVID-19 Lineage Assigner (https://pangolin.cog-uk.io/, last accessed 4/18/22).

### SARS-CoV-2 Variant Panel Testing

We recovered residual viral RNA from all 391 specimens from -80°C storage to undergo testing on the Agena MassARRAY^®^ SARS-CoV-2 Variant Panel v3 (https://www.agenabio.com/products/panel/coronavirus-sars-cov-2-variant-detection-research-panel/#:~:text=The%20MassARRAY%C2%AE%20SARS%2DCoV,SARS%2DCoV%2D2%20variants., last accessed 4/25/22). The panel combines RT-PCR and matrix-assisted laser desorption/ionization time-of-flight (MALDI-TOF) to detect targeted viral polymorphisms in the Spike (*S*) gene (**Fig. 1A**). It consists of a two-well multiplex qualitative assay that utilizes primer mixes that target a total of 36 polymorphisms which – in various signature combinations – reflect 16 distinct SARS-CoV-2 variants (**Fig. 1B**).

**FIG 1.**
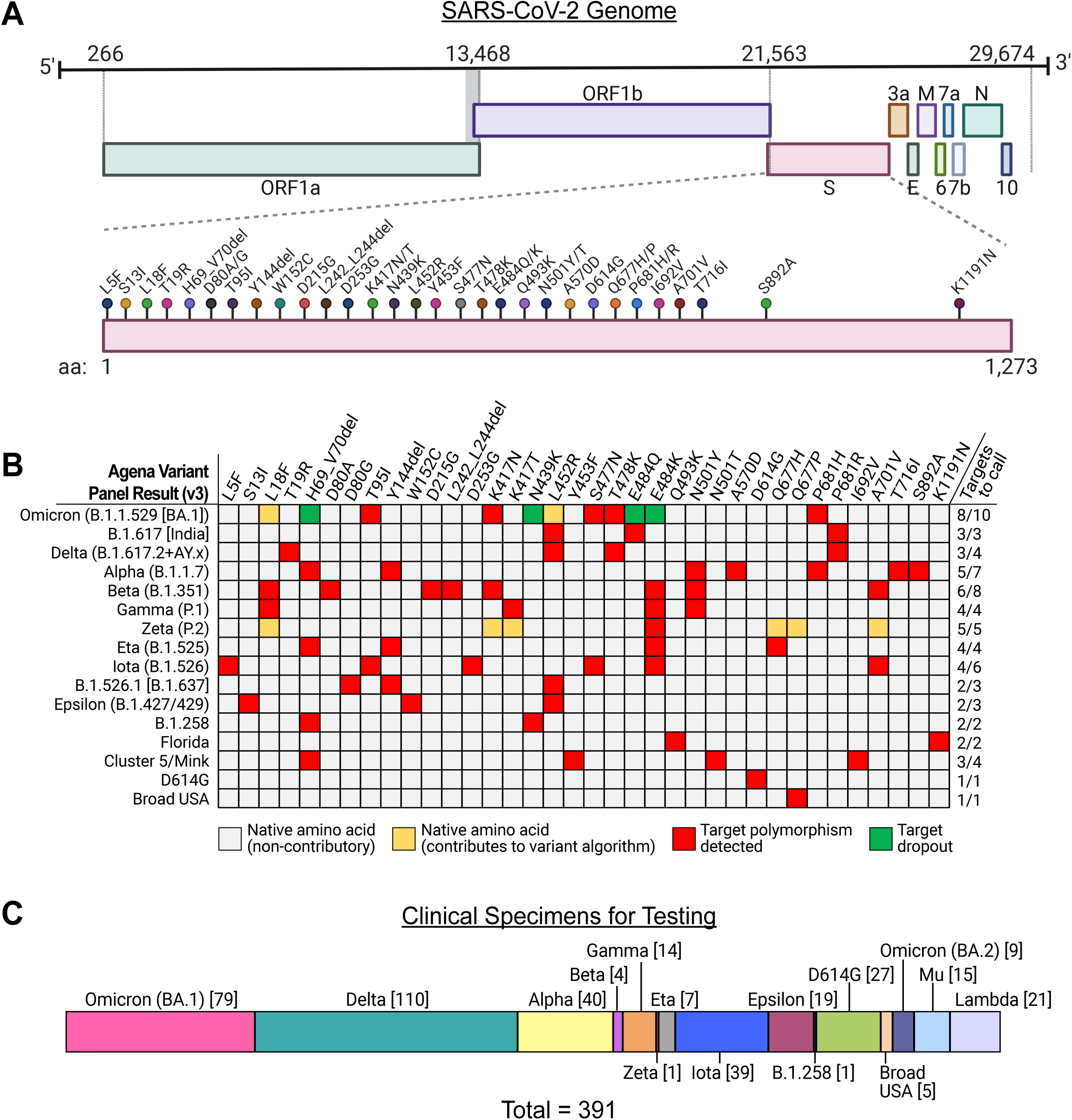
Detection of viral variants by the Agena MassARRAY^®^ SARS-CoV-2 Variant Panel. (A) SARS-CoV-2 genome with nucleotide positions from 5’-to-3’ direction depicted above. *S* gene polymorphisms targeted by the variant panel (lollipops) and corresponding amino acids are depicted below. (B) A color map depicts algorithms of target combinations that define 16 distinct SARS-CoV-2 variants on the panel. Variant results are depicted (left) which include the WHO designation (e.g., Omicron, Delta, etc.) and corresponding PANGO lineage assignments. Note that the B.1.526.1 variant was re-designated as B.1.637 to distinguish it from the Iota variant lineage (https://cov-lineages.org/lineage_list.html, last accessed 4/26/22). The minimum number of targets required to support the corresponding variant result are indicated (right). Target results are depicted as colored cells indicating detectable native (e.g., unchanged from Wuhan-Hu-1 reference) amino acids which do not contribute to the variant target algorithm (grey), detectable native amino acids which do contribute to the algorithm (yellow), detectable amino acid polymorphisms (red), and dropout of the given target polymorphism. (C) Phylogenetic composition of 391 clinical specimen viral RNA recovered for diagnostic evaluation of the variant panel. Numbers of each lineage tested are depicted in brackets.

#### RT-PCR and generation of analytes

Per manufacturer’s protocol, for each specimen, viral RNA underwent RT-PCR by combining 0.355μL nuclease-free water, 1μL RT-PCR Mastermix, 0.125μL RNase Inhibitor, and 0.020μL of MMLV Enzyme in each of two wells in a 384-well format. To one well, 0.500μL SARS-CoV-2 Variant v3 PCR Primer P01 was added, and to the second, 0.500μL SARS-CoV-2 Variant v3 PCR Primer P02 was added. Three microliters of sample RNA were added to each of the two wells for a final RT-PCR reaction volume of 5μL. Four positive controls of synthetic SARS-CoV-2 RNA (Twist Synthetic SARS-CoV-2 RNA Controls 1 (MT007544.1, #102019), 14 (B.1.1.7_710528, #103907), 16 (EPI_ISL_678597, #104043), and 17 (EPI_ISL_792683, #104044)) were diluted in a mixture of nuclease-free water (Ambion #AM9916) and human liver total RNA (Takara Bio #636531) and included in each RT-PCR run. This resulted in a total of 8 wells with 1,500 SARS-CoV-2 genome copies/well and 10ng human liver total RNA/well for all positive controls. A negative control of nuclease-free water was included in each RT-PCR run. RT-PCR thermocycler conditions are depicted in **Table S1**.

RT-PCR products underwent reaction with shrimp alkaline phosphatase (SAP). Directly to each RT-PCR well, a mastermix of 1.53μL nuclease-free water, 0.17μL SAP Buffer, and 0.30μL SAP was added for a total volume of 7μL including 2μL of SAP mastermix. SAP reaction thermocycler conditions are described in **Table S2**.

Extension products were generated with SARS-CoV-2 Variant v3 Extend Primers using the iPLEX^®^ Pro Reagent Set. Mastermixes were created as follows: 1.06μL nuclease-free water, 0.20μL iPLEX Buffer Plus, GPR; 0.20μL iPLEX Termination Mix, 0.04μL iPLEX Pro Enzyme, and 0.50μL MassARRAY^®^ SARS-CoV-2 Variant v3 Extend Primers (E01 or E02. Two microliters of E01 mastermix were added to each well amplified by P01 primers, and 2μL E02 mastermix was added to each well amplified by P02 primers for the total extension reaction volume of 9μL. Extension thermocycler conditions are detailed in **Table S3**.

#### Analyte dispensing, data acquisition, and data analyses

Twenty microliters of nuclease-free water were added to each well prior to desalting and dispensing in the MassARRAY^®^ System for data acquisition. Analytes were desalted using suspended clean resin (Agena #08060) and dispensed onto SpectroCHIP^®^ Arrays (CPM-384) for data acquisition with the MassARRAY^®^ Analyzer with Chip Prep Module 384 as per manufacturer’s protocol. Instrument settings for iPLEX Pro genotyping panels were used with Genotype+Area selected as the Process Method. After data acquisition, MassARRAY^®^ Typer Analyzer was used to analyze data and generate variant report output results for each specimen. All variant target results and individual target results for each specimen are depicted in **Table S4**.

### Diagnostic performance analyses

To compare performance of the panel to the WGS “gold standard”, we generated 2×2 contingency tables for detected and not detected results of each variant call or target call. To measure the level of agreement between WGS and the variant panel, we performed agreement analyses with kappa (κ) results and 95% confidence intervals (95% CI) using the publicly-available GraphPad Prism web calculator (https://graphpad.com/quickcalcs/kappa2/, last accessed April 20, 2022). Level of agreement was interpreted from kappa values as previously described (39). Interpretations included no (κ < 0), slight (0 ≤ κ ≤ 0.20), fair (0.21 ≤ κ ≤ 0.40), moderate (0.41 ≤ κ ≤ 0.60), substantial (0.61 ≤ κ ≤ 0.80), and almost perfect agreement (0.80 ≤ κ ≤1.00). In addition, we measured diagnostic sensitivity and specificity and negative (NPV) and positive predictive values (PPV) for each variant and individual target polymorphism tested (GraphPad Prism v.9.3.1). Ninety-five percent confidence intervals were calculated by the hybrid Wilson/Brown method. Fisher’s exact tests were performed for each contingency table for each variant and/or target tested.

Statistical analyses were performed for variant call results of 12 of the possible 16 variants on the panel. We also performed these analyses for 30 of the 36 possible targets on the panel as clinical specimens that encoded 6 specific amino acid polymorphisms (D80G, Y453F, E484Q, Q493K, N501T, I692V) were not recovered for this study. In addition, we did not recover any specimens that harbored the native D614 amino acid (A23403 nucleotide), and, therefore, we were not able to compute level of agreement or diagnostic specificity for the D614G variant or D614G target calls. In addition, for performance analyses of targets H69_V70del, N439K, and E484K, we excluded specimens that resulted in target dropout as we could not infer nucleotide polymorphisms that caused dropout given that primer/probe sequences are proprietary and not known.

To assess prevalence of variant panel targets across Omicron sublineages, we interrogated publicly-available SARS-CoV-2 genomes on the GISAID database (last accessed May 6, 2022). Using the online graphical user interface, we counted the number of genomes that harbored each of the 36 possible substitutions for each of four Omicron sublineages: BA.2.12.1 (n = 12324 genomes), BA.3 (n = 184), BA.4 (n = 857), BA.5 (n = 437). Prevalence of each substitution was determined by dividing the number of genomes with the substitution by the total number of genomes analyzed for the sublineage in question.

### Display Items

All figures are original and were generated using the GraphPad Prism software, Microsoft Excel v16.60, and finished in Adobe Illustrator (v.26.1). **Fig. 1A** was created in BioRender.com and finished in Adobe Illustrator.

### Data availability

All single variant consensus genome sequences were deposited in the publicly-available Global Initiative on Sharing Avian Influenza Data (GISAID) database (www.gisaid.org) [accession identifiers indicated in **Table S4**]. The remaining 10 genomes with mixed patters of mutations were not deposited into GISAID as single variant consensus genomes could not be resolved (data available upon request).

## Results

We analyzed a diverse set of 391 SARS-CoV-2 viral RNA specimens that were collected from infected patients over 18 months of the COVID-19 pandemic in the NYC metropolitan area and Colombia. These RNA all underwent WGS that resulted in consensus genomes that comprise 56 distinct phylogenetic Pango lineages and corresponded to 12 of the 16 possible variant calls on the panel (**Fig. 1C**). These included 39 Iota (B.1.526), 40 Alpha (B.1.1.7), 110 Delta (B.1.617.2 (n = 3) + AY.x (n = 107)), and 79 Omicron (B.1.1.529 [BA.1 sublineage]) specimens. We also included 45 specimens that corresponded to 3 variants that are not defined by the panel (e.g., Lambda (C.37), Mu (B.1.621), Omicron (BA.2 sublineage)) to interrogate the assay’s ability to distinguish these based on target result patterns.

### Diagnostic performance of variant calling

To evaluate the diagnostic performance of the Agena MassARRAY^®^ Variant Panel, phylogenetic results of consensus sequences based on WGS data served as the “gold standard” for comparison. Of 391 specimens tested on the variant panel, there were 62 with variant calls that were discordant from WGS data; however, 45 of these consisted of specimens whose sequenced variant was without an appropriately defined variant algorithm on the panel. Therefore, only 17 (4.91%) of clinical RNA tested yielded discordant results between WGS and the expected results on the variant panel.

We measured the level of agreement between the overall variant calls from WGS and the variant panel (**Table 1**). Of the 12 panel variants in our study set, we performed agreement analyses on 11. We could not measure level of agreement – or diagnostic sensitivity/specificity – for the isolated D614G result because specimens which concurrently encoded the native D614 amino acid and did not yield any other variant result were not recovered for this study.

**TABLE 1.** Diagnostic agreement between WGS and panel variant calls.

| Variant | Number of Specimens<sup>a</sup> | Agreement (%) | Kappa (95% CI) | Interpretation<sup>b</sup> |
| --- | --- | --- | --- | --- |
| Omicron (BA.1) | 79 | 98.72 % | 0.959 (0.924-0.995) | Almost perfect |
| Delta | 110 | 98.72 % | 0.968 (0.940-0.996) | Almost perfect |
| Alpha | 40 | 100.00 % | 1.000 (1.000-1.000) | Almost perfect |
| Beta | 4 | 99.74 % | 0.856 (0.577-1.000) | Almost perfect |
| Gamma | 14 | 99.74 % | 0.964 (0.894-1.000) | Almost perfect |
| Zeta | 1 | 95.91 % | -0.005 (-0.014-0.004) | None |
| Eta | 7 | 98.47 % | 0.247 (0.147-0.640) | Fair |
| Iota | 39 | 99.74 % | 0.986 (0.957-1.000) | Almost perfect |
| Epsilon | 19 | 100.00 % | 1.000 (1.000-1.000) | Almost perfect |
| B.1.258 | 1 | 100.00 % | 1.000 (1.000-1.000) | Almost perfect |
| D614G<sup>c</sup> | 27 | NA | NA | NA |
| Broad USA | 5 | 100.00 % | 1.000 (1.000-1.000) | Almost perfect |

<sup>a</sup> Number of specimens confirmed by WGS as the indicated variant.
<sup>b</sup> Interpretation of level of agreement is based on reference (39).
<sup>c</sup> Agreement analyses were not performed as specimens that harbor the native D614 amino acid were not recovered for testing. NA, not available.

Overall, we observed a high level of agreement (e.g., κ ≥ 0.856) for 9/11 variants including the contemporary Delta (B.1.617.2+AY.x) and Omicron (BA.1) variants. The Zeta (P.2) and Eta (B.1.525) variant calls demonstrated the lowest level of agreement. The single Zeta variant confirmed by WGS that was tested (PV26936) resulted as the Florida variant on the panel (**Table S4**). Interestingly, although the E484K and the four native amino acids – L18, K417, A701, Q677H – were correctly detected as part of the Zeta target algorithm, detection of K1191N and Q493K targets met the Florida variant target result criteria. The low level of agreement for the Zeta variant call is also impacted by the fact that all 15 Mu (B.1.621) specimens tested on the panel were incorrectly identified as Zeta; however, it is important to note that the Mu variant is not yet defined on this panel. Of the 7 Eta (B.1.525) variants tested on the panel, only 1 correctly identified as Eta while the remaining 6 resulted as detected D614G. These did not meet the minimum number of detectable targets to yield the Eta result and only resulted in 2-3 of the 4 minimum required targets. These results may be the consequences of RNA degradation over long-term storage. For example, the six discordant specimens encode the H69_V70del by WGS, but all yielded dropout of that target on the panel which further supports this scenario.

We also measured the diagnostic sensitivity and specificity of the panel (**Fig. 2A-B**) for the variants tested. Diagnostic sensitivity ranged from 0% (95% CI: 0-94.87%) [Zeta] to 100% (95% CI: 91.24-100%) [Alpha]. Unsurprisingly, the specimens for which we had limited (e.g., <10) specimens available for testing had the lowest measured sensitivities and broadest CIs including Zeta and Eta variants. Among variants for which we recovered >10 specimens for testing, diagnostic sensitivity was ≥93.67% [Omicron (BA.1)]. In addition, the variant panel demonstrated a high level of diagnostic specificity across all 11 variants tested that ranged from 96.15% (95% CI: 93.75-97.66%) for the Zeta variant to >99% for all other variants (**Fig. 2B**). Furthermore, excluding the Zeta variant, the panel results showed high PPVs (≥0.933) and high NPVs (≥0.984) for all variant calls (**Table S5**).

**FIG 2.**
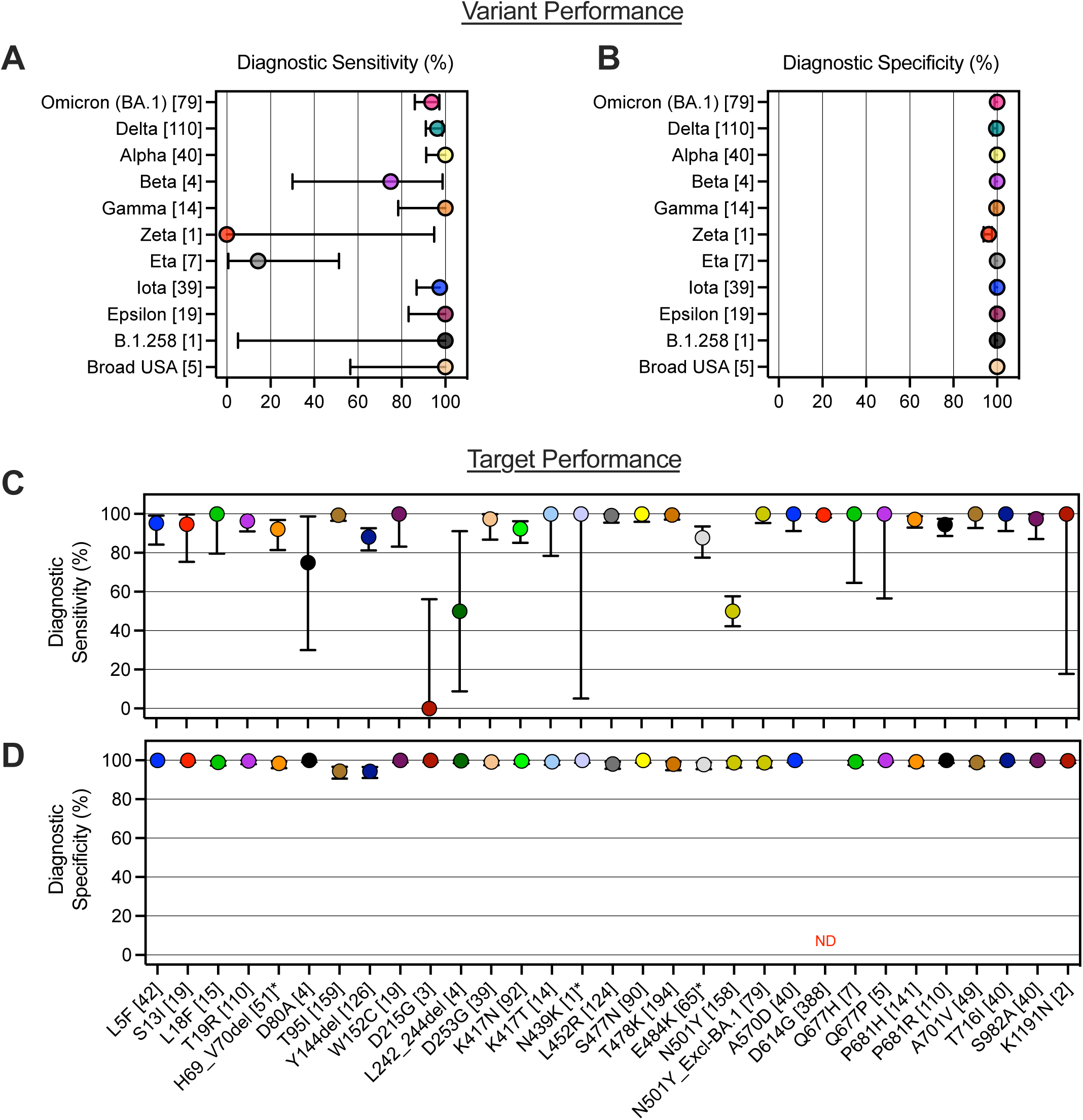
Diagnostic sensitivity and specificity of the Agena MassARRAY^®^ SARS-CoV-2 Variant Panel. (A) Diagnostic sensitivity and (B) diagnostic specificity of eleven variant calls on the panel are depicted. The number of specimens that correspond with each variant according to WGS are annotated in brackets. Depiction of (C) diagnostic sensitivity and (D) diagnostic specificity of each of thirty distinct panel targets. The number of specimens that correspond with each amino acid polymorphism according to WGS are annotated in brackets for each target. Asterisks (*) indicate targets for which dropout results were excluded from analyses (see Methods). For target N501Y, a separate diagnostic analysis was conducted excluding BA.1 specimens (“N501Y_Excl-BA.1”). Error bars reflect 95% CI in all four panels. ND, not determined.

### Diagnostic performance of distinct target calls

To evaluate the diagnostic capabilities of each of the 36 targeted polymorphisms that comprise the variant panel, we performed interrater agreement analyses and measured the diagnostic sensitivity and specificity of each assay target. Across all 391 viral RNA specimens, each of 30 of the possible 36 polymorphisms were present in at least one specimen.

When we performed agreement analyses on each of these 30 targets (**Table 2**), 25 demonstrated almost perfect levels of agreement (κ ≥ 0.820). The targets with suboptimal levels of agreement for our dataset included D215G (κ = 0; no agreement), L242_244del (κ = 0.568; moderate agreement), N501Y (κ = 0.528; moderate agreement), and K1191N (κ = 0.799; substantial agreement). This low level of agreement may be impacted by small sample sizes tested. Indeed, we only recovered 2-4 specimens that encoded each of the D215G, L242_244del, and K1191N targets, Therefore, small frequencies (e.g., 2-4) of inaccurate calls may explain this result. It is important to note that our study set did not include specimens with the native D614 amino acid (A24303 nucleotide), and level of agreement could not be calculated for the D614G target.

**TABLE 2.**
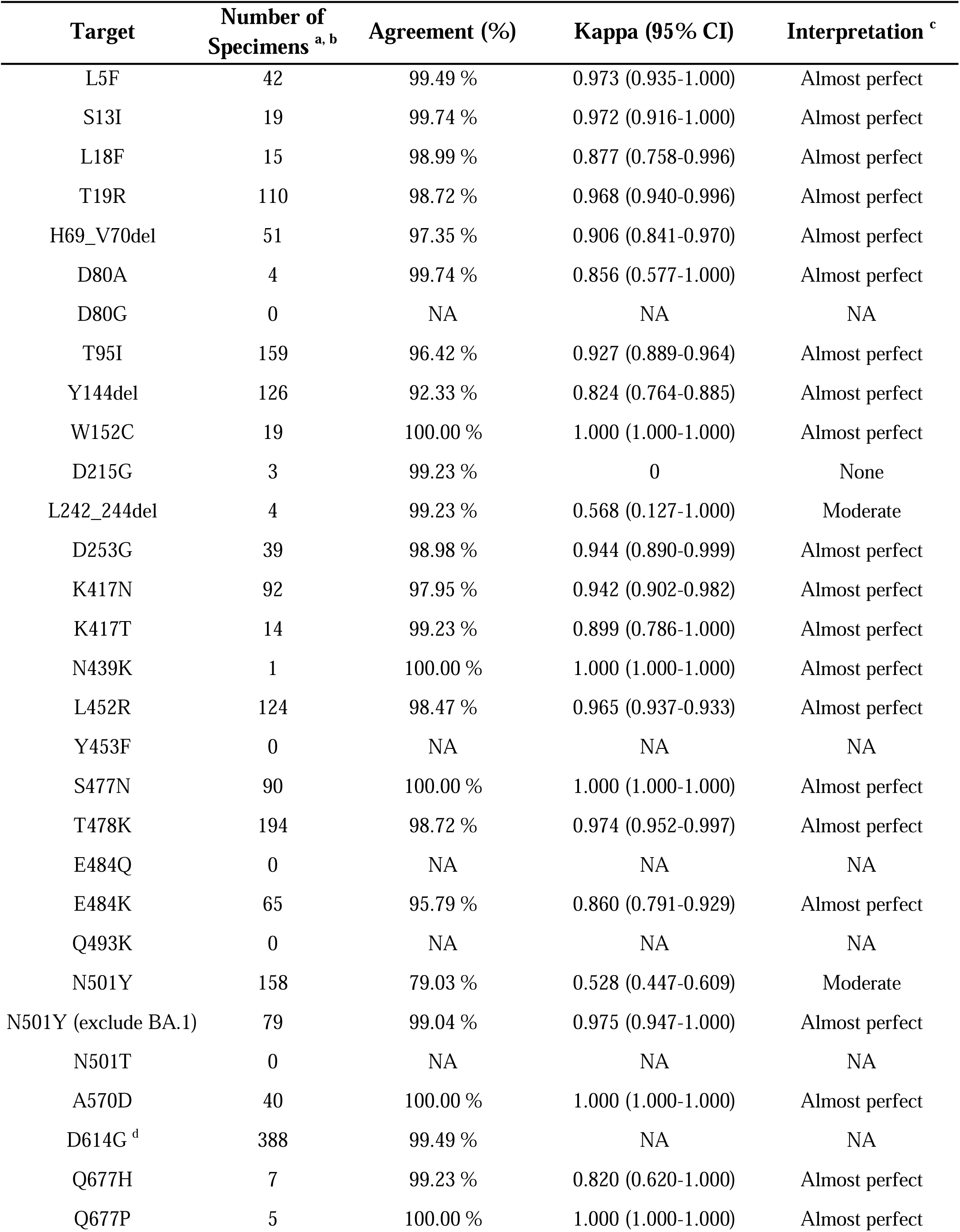
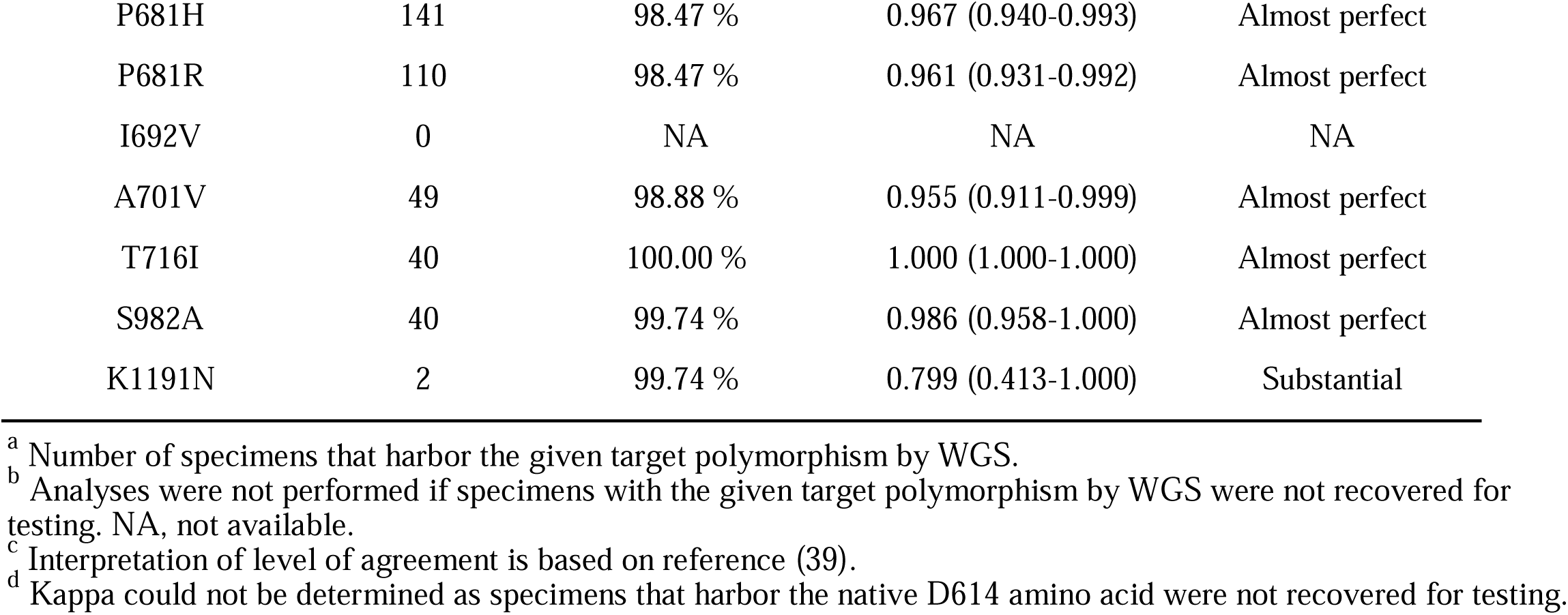
Diagnostic agreement between WGS and panel target calls.

Interestingly, for N501Y target, we found that of 158 specimens with the polymorphism by WGS, 79 yielded a false-negative result on the variant panel. All 79 belong to the Omicron (BA.1) variant lineage, and when reanalyzed excluding these BA.1 specimens, interrater agreement was almost perfect (κ = 0.975) (**Table 2**). This suggests genomic variation outside original assay design may impact primer/probe binding and yield distinct target results for novel variants.

Across the 30 targets tested, the average diagnostic sensitivity measured was 90.2% (**Fig. 2C**). The targets with the lowest sensitivities included D80A (75.00%, 95% CI: 30.06-98.72%), D215G (0%, 95% CI: 0.00-56.00%), L242_244del (50.00%, 95% CI: 9.00-91.00%), and N501Y (50.00%, 95% CI: 42.00-58.00%). Notably, when all BA.1 specimens were excluded from the analyses, the sensitivity of the N501Y target improved to 100.00% (95% CI: 95.00-100.00%).

The variant panel assay demonstrated a high diagnostic specificity across nearly all 30 targets tested in this study (**Fig. 2D**). On average, the diagnostic specificity was >99.00% across all of the tested diagnostic targets. Specificity was not calculated for the D614G target because no clinical specimens with the native D614 amino acid were recovered in this study. In addition, across the 30 targets, the PPVs and NPVs were 0.900 and 0.959, respectively (**Table S6**).

### Diagnostic target signatures of undefined variants

Given that the variant panel has a uniquely high level of multiplexing, we also interrogated the capabilities of the assay to reveal unique signatures of variants not defined by the panel software. To do this, we included 45 clinical specimens that included the older Lambda (C.37) (n = 21) and Mu (B.1.621) (n = 15) variants as well as the contemporary Omicron BA.2 variants (n = 9) recently captured in NYC (**Fig. 3A**). Each of the three were called as D614G, Zeta (P.2), and D614G, respectively on the panel.

**FIG 3.**
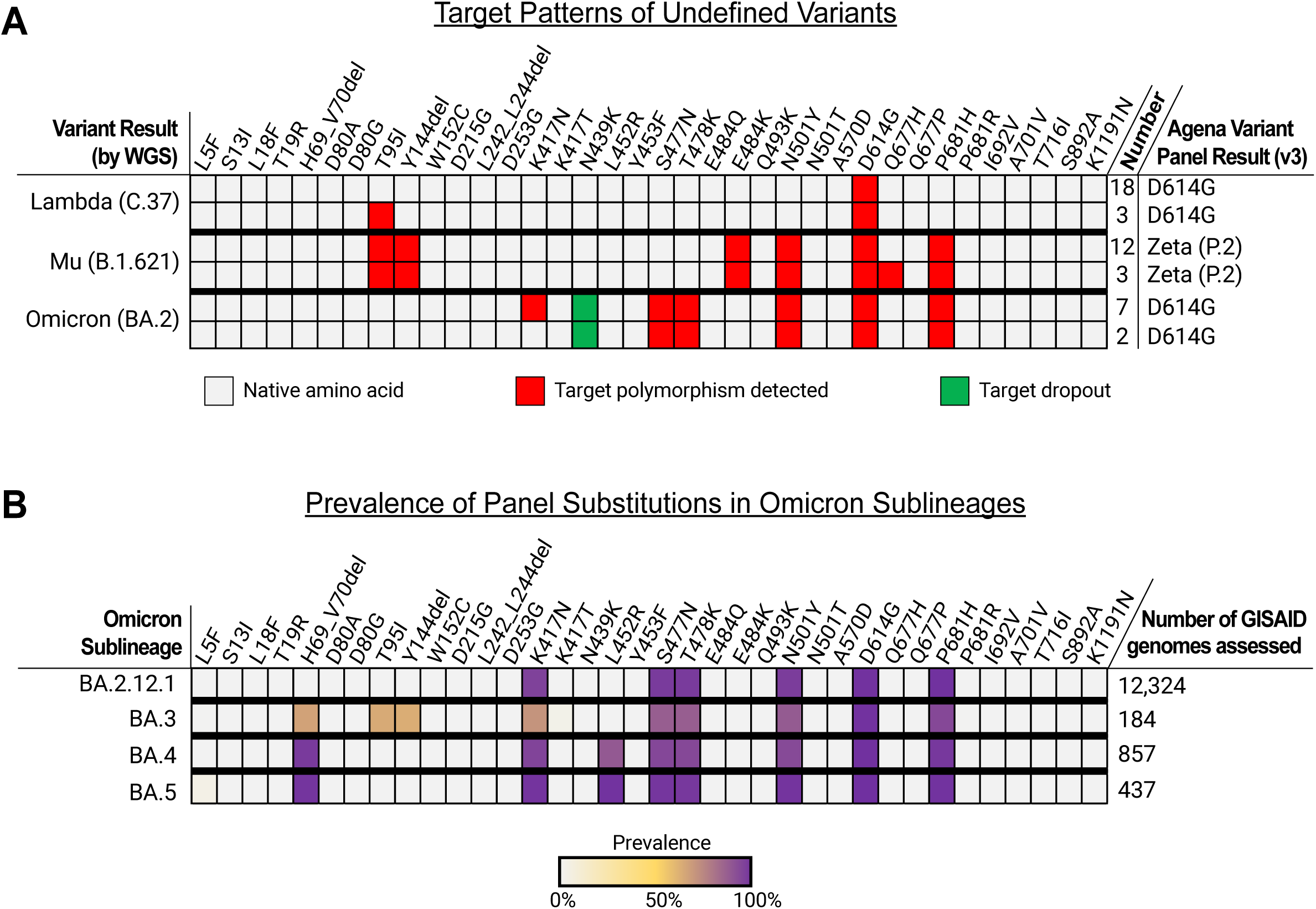
Target result patterns of undefined variants on the Agena MassARRAY^®^ SARS-CoV-2 Variant Panel. **(A)** A color map depicts the observed target results for three undefined SARS-CoV-2 variants tested on the panel: Lambda (C.37), Mu (B.1.621), and Omicron (BA.2). Distinct target patterns observed among each of the variant types are depicted. Cells indicate the distinct target results including detectable native amino acid (grey), detection of target polymorphism (red), and target dropout (green). The number of specimens that yielded each of the distinct target result patterns are indicated on the right as well as the output variant ID result generated by the variant panel software. (B) A heatmap depicts the measured prevalence of each variant panel target substitution among publicly-available Omicron sublineage genomes as of May 6, 2022.

Based on the current design of the panel, most of the Lambda specimens tested (18/21) only have detectable D614G polymorphisms among the 36 targets. The remaining 3 additionally yield a T95I (C21846U) polymorphism; however, this substitution is not found in any of the 3 consensus sequences, and, therefore, may represent a nonspecific reaction or a minority intra-host variant. Notably, this polymorphism is rare and found in only 24/10186 (0.23%) Lambda genomes deposited in GISAID (last accessed May 6, 2022). The current target design does not target common Lambda substitutions including G75V, T76I, D253N, L452Q, T859N, or deletions (e.g., amino acids 246-252).

All 15 Mu specimens were appropriately called as Zeta (P.2) based on the presence of the native L18, K417, Q677, and A701 amino acids as well as the E484K which meets the threshold for the Zeta variant call. All Mu specimens display a signature of six detectable targets that is unique among all other variant patterns on this assay: T95I, Y144del, E484K, N501Y, D614G, and P681H. Interestingly, none of the specimens’ consensus genome sequences encode the Y144del which suggests other sequence variation may alter primer/probe binding to this target. In fact, 12 of the Mu sequences harbor substitutions at positions 144-145 (e.g., Y144S, Y145N) which may impact target detection. The five other amino acid substitutions are encoded in the consensus genomes of all 15 specimens. Of note, although specimen K42 features an adenosine insertion at genome position 21995, the downstream nucleotide sequence encodes these amino acid polymorphisms. In addition to these 5 substitutions, there are 3 Mu specimens which yielded a detectable Q677H target. However, these genomes harbor the G23593 nucleotide which encodes the native Q677 amino acid and suggests this is a nonspecific result or detection of a minor intra-host variant.

We also found that the 9 Omicron BA.2 specimens generated a distinct target result signature on the variant panel assay. All 9 resulted in detection of S477N, T478K, N501Y, D614G, and P681H targets as well as dropout of the N439K target. The five detected targets each were confirmed by the presence of the amino acid substitutions in WGS data. We cannot delineate the cause of the N439K target dropout because we do not know primer/probe sequences at the site of the targeted nucleotide substitution at position 22879. However, one can speculate that sequence variation around this region may interfere with primer/probe binding. Indeed, all 9 of the BA.2 specimens harbor the T22882G polymorphism which results in the N440K substitution. Furthermore, the K417N substitution is found in all BA.2 consensus genomes but is only detected in 7 specimens. This may be the result of different nucleic acid quantities across specimens and reflect a limit in analytic sensitivity for the target. Together, these are important findings to gauge the capabilities of this assay to highlight unique target signatures of variants that may be captured by this platform.

Finally, to assess the capabilities of the assay to detect other Omicron sublineages (e.g., BA.2.12.1, BA.3, BA.4, BA.5), we independently interrogated the prevalence of each variant panel target substitution among publicly-available genomes (GISAID) (**Fig. 3B**). We found that 94.9-99.7% of BA.2.12.1 genomes harbor each of the K417N, S477N, T478K, N501Y, D614G, and P681H substitutions. With the exception of the N439K target dropout in BA.2 genomes tested in this study, the BA.2.12.1 target signature harbors the same detectable polymorphisms. While 70.7-99.5% of BA.3 genomes also encode the S477N, T478, N501Y, D614G, and P681H, 63.0-66.3% harbor the H69_V70del, T95I, and Y144del substitution which help to distinguish this sublineage from BA.2 and BA.2.12.1. Furthermore, 88.1% of BA.4 and 99.3% of BA.5 genomes encode the L452R substitution which may help to differentiate these genomes from other Omicron sublineages.

## Discussion

Monoclonal antibody treatments are effective in limiting severe COVID-19 but emerging variants of concern often carry mutations that render the virus partially or completely resistant to antibody neutralization (13, 14, 40–44). Rapid SARS-CoV-2 variant calling is therefore essential for personalized COVID-19 treatment interventions. With the advent of commercial and lab-developed variant panels, however, accurate variant requires robust, high-resolution platforms which are limited in number and have not undergone evaluation prior to implementation. Indeed, a multi-laboratory external quality assessment in late 2021 revealed gaps in calling of contemporary variants that stemmed from inadequate selection of diagnostic targets to discern between variants (27). Given this, highly multiplexed, efficient platforms are invaluable but are limited in number and warrant comprehensive evaluation before implementation in the molecular microbiology laboratory.

Here, we report a comprehensive diagnostic evaluation of one of the highest multiplexed variant panel assays on the market. Based on a diverse cohort of clinical specimens across two continents and a wide timeline of the pandemic, we highlight almost perfect levels of interrater agreement between this assay and the “gold standard” WGS for 9 of 11 variants and 25 of 30 distinct targets tested. The assay has a high diagnostic specificity across all variants (≥96.15%) and all targets (≥94.34%) tested. Furthermore, the panel shows a high diagnostic specificity and sensitivity for contemporary variants in global circulation (e.g., Delta, Omicron (BA.1)).

Our study does present some limitations particularly with respect to limited sampling. While the panel has defined target signatures for 16 different variants, we were only able to recover clinical specimens that corresponded to 11 of these variants for testing. Indeed, variants with the lowest level of agreement and diagnostic performance metrics were those with some of the fewest specimens recovered and tested (e.g., Zeta (n = 1), Beta (n = 4), Eta (n = 7). We also did not include specimens from the early phase of the pandemic including D614 viruses (45, 46) which limited diagnostic analyses of the D614G variant and individual target. It is important to note, however, that the D614G polymorphism has undergone positive selection to eventuate emergent variants (47), and these older viruses have largely been replaced by the emergent Omicron lineage(s) (6, 48). We also recognize that we did not conduct this study at the extraction step of clinical specimens given limited availability of remnant upper respiratory or saliva specimens.

A unique benefit of a highly multiplexed molecular assay is its adaptability to the natural evolution of the pathogen at hand which confers the ability to identify changes in circulating viruses that manifest as distinct target result signatures. To assess this potential, we included undefined variants to determine if the discrete assay target result patterns could elucidate a variant’s identity without necessarily providing a defined result as the current software stands. Testing of Mu specimens resulted in a distinguishable combination of 5 detectable substitutions, but each result was interpreted as a Zeta (P.2) variant. This scenario highlights the utility of distinct target results to point to new viruses that rapidly arise the circulating milieu of variants. Moreover, this underscores the importance of adaptable target result interpretation software to address acute changes detected in patient populations. We also tested clinical specimens that corresponded with the most current variant in circulation – the Omicron sublineage BA.2 – which has largely replaced BA.1 globally over January through April 2022 (https://covariants.org/, last accessed 4/26/22). From our results, we report a BA.2-specific pattern of target results on this panel that can be used to readily discriminate the BA.2 from the BA.1 subtype. The following weeks/months will be key to monitoring to understand the utility of this platform for capturing other emerging Omicron sublineages (e.g., BA.2.12.1, BA.3, BA.4, BA.5).

Accurate identification of currently circulating and emerging SARS-CoV-2 variants is key to effective pathogen surveillance and providing optimal care to patients. Although WGS is the mainstay for surveillance, it is important to consider in the global context of a pandemic, this may not be a realistic technology for many LICs and LMCs (29). Therefore, there is a great need for rapid, cost-effective, conventional technologies (e.g., RT-PCR) in the clinical laboratory. However, these require increased diagnostic resolution to adequately capture viral evolution and meet the needs of pathogen surveillance. Thus, highly multiplexed molecular assays such as the one presented benefit from high discriminatory power and are a vital tool to shed light on changing viral dynamics.

## Data Availability

All single variant consensus genome sequences were deposited in the publicly-available Global Initiative on Sharing Avian Influenza Data (GISAID) database (www.gisaid.org) [accession identifiers indicated in Table S4]. The remaining 10 genomes with mixed patters of mutations were not deposited into GISAID as single variant consensus genomes could not be resolved (data available upon request).

## Acknowledgments

We thank the members of MSHS MML, Simon, and van Bakel laboratories. We are grateful for the continuous expert guidance provided by the ISMMS Program for the Protection of Human Subjects (PPHS).

We are thankful for the efforts of the Mount Sinai PSP Study Group members (in alphabetical order): Hala Alshammary, Angela A. Amoako, Mahmoud H. Awawda, Christian Cognigni, Aria Rooker, Jose Polanco, Levy A. Sominsky, Komal Srivastava, Paras Shrestha, Steve Shi, Jian Zhang, Zain Khalil.

## Author contributions

M.M.H., R.B., P.S., M.S.PSP.S.G., A.E.PM., M.R.G., M.D.N., M.M., N.L., A.R., S.C., N.B., J.D.R. and E.M.S. provided clinical samples for the study. M.M.H., R.B., P.S., M.S.PSP.S.G., and A.E.PM. accessioned clinical samples. A.S.GR., A.v.d.G, K.F., M.S.PSP.S.G., M.M., N.L., A.R., S.A.C., N.B., L.H.P., and H.v.B. performed viral genome sequencing experiments. R.S. provided NGS services. A.S.GR. and M.S.PSP.S.G. performed genome assembly and data curation. M.M.H., R.B., P.S., J.D.R., M.R.G., M.D.N., C.C.C., V.S., H.v.B., E.M.S., and A.E.PM analyzed, interpreted, or discussed data. M.M.H. wrote the manuscript. M.M.H., J.D.R., and A.E.PM. conceived the study. E.M.S. and A.E.PM. supervised the study. H.v.B., V.S., A.E.PM., and E.M.S. raised financial support.

M.M.H. and A.E.PM are the guarantors of this work and, as such, had full access to all of the data in the study and take responsibility for the integrity of the data and the accuracy of the data analysis.

