## Supplemental Tables for "A robust, highly multiplexed mass spectrometry assay to identify SARS-CoV-2 variants"

**SUPPLEMENTAL INFORMATION**

**Table S1. RT-PCR thermocycling conditions**

| **Step** | **Description** | **Temperature** | **Time** | **Cycles** |
| --- | --- | --- | --- | --- |
| 1 | Uracil-DNA-glycosylase | 25°C | 5 minutes | 1 |
| 2 | RNA reverse-transcription | 50°C | 10 minutes | 1 |
| 3 | Polymerase activation | 95°C | 2 minutes | 1 |
| 4 | PCR | 95°C | 5 seconds | 10 |
| 5 |  | 65°C (-1°C /cycle) | 10 seconds |  |
| 6 |  | 72°C | 5 seconds |  |
| 7 |  | 95°C | 5 seconds | 35 |
| 8 |  | 55°C | 10 seconds |  |
| 9 |  | 72°C | 5 seconds |  |
| 10 | Final extension | 72°C | 5 minutes | 1 |
| 11 | Sample preservation | 10°C | --- | Hold |

**Table S2. SAP reaction conditions**

| **Step** | **Description** | **Temperature** | **Time** | **Cycles** |
| --- | --- | --- | --- | --- |
| 1 | Dephosphorylation | 37°C | 10 minutes | 1 |
| 2 | Enzyme inactivation | 85°C | 5 minutes | 1 |
| 3 | Sample preservation | 10°C | --- | Hold |

**Table S3. Extension thermocycler conditions**

| **Step** | **Description** | **Temperature** | **Time** | **Sub-Cycles** | **Cycles** |
| --- | --- | --- | --- | --- | --- |
| 1 | Initial denaturation | 95°C | 30 seconds | 1 |  |
| 2 | Denaturation | 95°C | 5 seconds | 1 | 40 |
| 3 | Annealing/Extension | 52°C | 5 seconds | 5 |  |
| 4 | Denaturation | 80°C | 5 seconds |  |  |
| 5 | Final extension | 72°C | 3 minutes | 1 |  |
| 6 | Sample preservation | 10°C | --- | Hold |  |

**Table S4. Agena MassARRAY^®^ SARS-CoV-2 Variant Panel v3 (RUO) results**

| **ID** | **Collection Date** | **GISAID Accession ^a^** | **PANGO Lineage (WGS)** | **Agena Variant Results ^b^** | **Genetic Markers Detected ^c^** |
| --- | --- | --- | --- | --- | --- |
| PV17741 | 9/2/20 | EPI_ISL_802220 | B.1.239 | D614G Detected. | D614G \| K417T \| Q493K |
| PV17750 | 9/3/20 | EPI_ISL_802222 | B.1.1.434 | D614G Detected. | D614G |
| PV17762 | 9/3/20 | EPI_ISL_801862 | B.1.1.50 | D614G Detected. | D614G \| K417T |
| PV17911 | 9/3/20 | EPI_ISL_802223 | B.1.1.1 | D614G Detected. | D614G |
| PV17731 | 9/4/20 | EPI_ISL_801922 | B.1.240 | D614G Detected. | D614G |
| PV17864 | 9/4/20 | EPI_ISL_802225 | B.1.1.432 | D614G Detected. | D614G \| H69_V70del_Dropout |
| PV17895 | 9/4/20 | EPI_ISL_801893 | B.1.2 | D614G Detected. | D614G |
| PV17896 | 9/4/20 | EPI_ISL_802226 | B.1.2 | D614G Detected. | D614G |
| PV17754 | 9/5/20 | EPI_ISL_802228 | B.1.2 | D614G Detected. | D614G \| D80G \| L18F \| N439K_Dropout \| Q493K |
| PV17758 | 9/5/20 | EPI_ISL_801905 | B.1.1.265 | D614G Detected. | D614G |
| PV17759 | 9/5/20 | EPI_ISL_801881 | B.1.240 | D614G Detected. | D614G |
| PV17761 | 9/5/20 | EPI_ISL_802229 | B.1.2 | D614G Detected. | D614G |
| PV19592 | 9/6/20 | EPI_ISL_801941 | B.1.1 | D614G Detected. | D614G |
| PV18393 | 9/7/20 | EPI_ISL_801926 | B.1.1.434 | D614G Detected. | D614G |
| PV17748 | 9/8/20 | EPI_ISL_801874 | B.1.369 | D614G Detected. | D614G |
| PV17846 | 9/8/20 | EPI_ISL_6491830 | B.1 | D614G Detected. | D614G |
| PV17879 | 9/8/20 | EPI_ISL_802230 | B.1.361 | D614G Detected. | D614G |
| PV17897 | 9/8/20 | EPI_ISL_802231 | B.1 | D614G Detected. | D614G |
| PV17921 | 9/8/20 | EPI_ISL_802232 | B.1.265 | D614G Detected. | D614G \| D80G \| H69_V70del_Dropout \| K417N \| L18F |
| PV17923 | 9/8/20 | EPI_ISL_802234 | B.1.1.434 | D614G Detected. | D614G |
| PV19595 | 9/8/20 | EPI_ISL_802235 | B.1.1 | D614G Detected. | D614G |
| PV17905 | 9/9/20 | EPI_ISL_802237 | B.1.240 | D614G Detected. | D614G |
| PV17924 | 9/9/20 | EPI_ISL_801940 | B.1.1 | D614G Detected. | D614G |
| PV17881 | 9/10/20 | EPI_ISL_801883 | B.1.1.231 | D614G Detected. | D614G |
| PV17882 | 9/10/20 | EPI_ISL_802238 | B.1.1.434 | D614G Detected. | D614G |
| PV18993 | 9/10/20 | EPI_ISL_802243 | B.1 | D614G Detected. | D614G |
| PV17858 | 9/11/20 | EPI_ISL_801900 | B.1.1 | D614G Detected. | D614G |
| PV19366 | 9/28/20 | EPI_ISL_801959 | B.1.258 | B.1.258 Detected. D614G Detected. | D614G \| D80G \| H69_V70del \| L18F \| N439K \| Q493K |
| PV26936 | 1/25/21 | EPI_ISL_1708896 | P.2 | Florida Detected. | E484K \| K1191N \| N439K_Dropout \| Q493K |
| PV27007 | 1/26/21 | EPI_ISL_1708926 | B.1.351 | Beta (B.1.351) Detected. D614G Detected. | A701V \| D614G \| D80A \| E484K \| K417N \| L242_L244del \| N501Y |
| PV27065 | 1/27/21 | EPI_ISL_1708944 | B.1.429 | B.1.427/B.1.429 (20C.Cal) Detected. D614G Detected. | D614G \| L452R \| S13I \| W152C |
| PV27047 | 1/28/21 | EPI_ISL_1300758 | B.1.526 | D614G Detected. Iota (B.1.526) Detected. | A701V \| D253G \| D614G \| E484K \| T95I |
| PV27056 | 1/29/21 | EPI_ISL_1300899 | B.1.526 | D614G Detected. Iota (B.1.526) Detected. | A701V \| D253G \| D614G \| E484K \| H69_V70del \| L5F \| T95I |
| PV27058 | 1/29/21 | EPI_ISL_1300900 | B.1.526 | D614G Detected. Iota (B.1.526) Detected. | A701V \| D253G \| D614G \| L5F \| S477N \| T95I |
| PV27103 | 1/29/21 | EPI_ISL_1708950 | B.1.2 | Broad USA Detected. D614G Detected. | D614G \| H69_V70del_Dropout \| Q677P |
| PV27081 | 1/30/21 | EPI_ISL_1301403 | B.1.429 | B.1.427/B.1.429 (20C.Cal) Detected. D614G Detected. | D614G \| L452R \| S13I \| W152C |
| PV27085 | 1/30/21 | EPI_ISL_1300902 | B.1.526 | D614G Detected. Iota (B.1.526) Detected. | A701V \| D253G \| D614G \| L5F \| T95I |
| PV27082 | 1/30/21 | EPI_ISL_1301404 | B.1.2 | Broad USA Detected. D614G Detected. | D614G \| E484-K/Q_Dropout \| H69_V70del_Dropout \| Q677P |
| PV27090 | 1/31/21 | EPI_ISL_5253374 | B.1.526 | D614G Detected. Iota (B.1.526) Detected. | A701V \| D253G \| D614G \| L5F \| T95I |
| PV27094 | 1/31/21 | EPI_ISL_1301407 | B.1.429 | B.1.427/B.1.429 (20C.Cal) Detected. D614G Detected. | D614G \| L452R \| W152C |
| PV27105 | 1/31/21 | EPI_ISL_1708951 | B.1.526 | D614G Detected. Iota (B.1.526) Detected. | A701V \| D253G \| D614G \| L5F \| T95I |
| PV27109 | 2/2/21 | EPI_ISL_1300804 | B.1.1.7 | Alpha (B.1.1.7) Detected. D614G Detected. | A570D \| D614G \| H69_V70del \| N501Y \| P681H \| S982A \| T716I \| Y144del_Composite |
| PV27111 | 2/2/21 | EPI_ISL_1300903 | B.1.526 | D614G Detected. Iota (B.1.526) Detected. | A701V \| D253G \| D614G \| L5F \| T95I |
| PV27533 | 2/2/21 | EPI_ISL_1708957 | B.1.1.7 | Alpha (B.1.1.7) Detected. D614G Detected. | A570D \| D614G \| H69_V70del \| N501Y \| P681H \| S982A \| T716I \| Y144del_Composite |
| PV27541 | 2/2/21 | EPI_ISL_1708956 | B.1.526 | D614G Detected. Iota (B.1.526) Detected. | A701V \| D253G \| D614G \| E484K \| L5F \| T95I |
| PV27526 | 2/3/21 | EPI_ISL_1300904 | B.1.526 | D614G Detected. Iota (B.1.526) Detected. | A701V \| D253G \| D614G \| L5F \| T95I |
| PV27528 | 2/3/21 | EPI_ISL_1300905 | B.1.526 | D614G Detected. Iota (B.1.526) Detected. | A701V \| D253G \| D614G \| E484K \| L5F \| T95I |
| PV27535 | 2/3/21 | EPI_ISL_1300907 | B.1.526 | D614G Detected. Iota (B.1.526) Detected. | A701V \| D253G \| D614G \| L5F \| T95I |
| PV27539 | 2/3/21 | EPI_ISL_1300908 | B.1.526 | D614G Detected. Iota (B.1.526) Detected. | A701V \| D253G \| D614G \| E484K \| L5F \| T95I |
| PV27534 | 2/3/21 | EPI_ISL_1300906 | B.1.526 | D614G Detected. Iota (B.1.526) Detected. | A701V \| D253G \| D614G \| E484K \| L5F \| T95I |
| PV27543 | 2/3/21 | EPI_ISL_1300768 | B.1.1.7 | Alpha (B.1.1.7) Detected. D614G Detected. | A570D \| D614G \| H69_V70del \| N501Y \| P681H \| S982A \| T716I \| Y144del_Composite |
| PV27547 | 2/4/21 | EPI_ISL_1301418 | B.1.427 | B.1.427/B.1.429 (20C.Cal) Detected. D614G Detected. | D614G \| L452R \| S13I \| W152C |
| PV27553 | 2/4/21 | EPI_ISL_1300809 | B.1.1.7 | Alpha (B.1.1.7) Detected. D614G Detected. | A570D \| D614G \| H69_V70del \| N501Y \| P681H \| S982A \| T716I \| Y144del_Composite |
| PV27549 | 2/4/21 | EPI_ISL_1300909 | B.1.526 | D614G Detected. Iota (B.1.526) Detected. | A701V \| D253G \| D614G \| E484K \| L5F \| T95I |
| PV27566 | 2/4/21 | EPI_ISL_5253375 | B.1.526 | D614G Detected. | A701V \| D614G \| T95I |
| PV27567 | 2/4/21 | EPI_ISL_5253376 | B.1.526 | D614G Detected. Iota (B.1.526) Detected. | A701V \| D253G \| D614G \| L5F \| T95I |
| PV27571 | 2/4/21 | EPI_ISL_5336550 | B.1.526 | D614G Detected. Iota (B.1.526) Detected. | A701V \| D253G \| D614G \| L5F \| T95I |
| PV27573 | 2/4/21 | EPI_ISL_1708964 | B.1.1.7 | Alpha (B.1.1.7) Detected. D614G Detected. | A570D \| D614G \| H69_V70del \| N501Y \| P681H \| S982A \| T716I \| Y144del_Composite |
| PV27597 | 2/4/21 | EPI_ISL_1708969 | B.1.526 | D614G Detected. Iota (B.1.526) Detected. | A701V \| D253G \| D614G \| E484K \| L5F \| T95I |
| PV27587 | 2/5/21 | EPI_ISL_1300910 | B.1.526 | D614G Detected. Iota (B.1.526) Detected. | A701V \| D253G \| D614G \| E484K \| L5F \| T95I |
| PV27584 | 2/5/21 | EPI_ISL_1301426 | B.1.427 | B.1.427/B.1.429 (20C.Cal) Detected. D614G Detected. | D614G \| L452R \| S13I \| W152C |
| PV27589 | 2/5/21 | EPI_ISL_1300912 | B.1.526 | D614G Detected. Iota (B.1.526) Detected. | A701V \| D253G \| D614G \| E484K \| L5F \| T95I |
| PV27590 | 2/5/21 | EPI_ISL_1300913 | B.1.526 | D614G Detected. Iota (B.1.526) Detected. | A701V \| D253G \| D614G \| E484K \| L5F \| T95I |
| PV27593 | 2/5/21 | EPI_ISL_1300914 | B.1.526 | D614G Detected. Iota (B.1.526) Detected. | A701V \| D253G \| D614G \| E484K \| L5F \| T95I |
| PV27608 | 2/6/21 | EPI_ISL_1300915 | B.1.526 | D614G Detected. Iota (B.1.526) Detected. | A701V \| D253G \| D614G \| E484K \| N439K_Dropout \| T95I |
| PV27609 | 2/6/21 | EPI_ISL_1300916 | B.1.526 | D614G Detected. Iota (B.1.526) Detected. | A701V \| D253G \| D614G \| E484K \| L5F \| T95I |
| PV27610 | 2/6/21 | EPI_ISL_1301429 | B.1.429 | B.1.427/B.1.429 (20C.Cal) Detected. D614G Detected. | D614G \| L452R \| S13I \| W152C |
| PV27616 | 2/7/21 | EPI_ISL_1300810 | B.1.1.7 | Alpha (B.1.1.7) Detected. D614G Detected. P.2 Detected. | A570D \| D614G \| E484K \| H69_V70del \| N501Y \| P681H \| S982A \| T716I \| Y144del_Composite |
| PV27618 | 2/7/21 | EPI_ISL_1300818 | B.1.526 | D614G Detected. Iota (B.1.526) Detected. | A701V \| D253G \| D614G \| E484K \| L5F \| T95I |
| PV27631 | 2/7/21 | EPI_ISL_1300805 | B.1.1.7 | Alpha (B.1.1.7) Detected. D614G Detected. | A570D \| D614G \| H69_V70del \| N501Y \| P681H \| S982A \| T716I \| Y144del_Composite |
| PV27651 | 2/7/21 | EPI_ISL_1708981 | B.1.526 | D614G Detected. Iota (B.1.526) Detected. P.2 Detected. | A701V \| D253G \| D614G \| E484K \| L5F \| S477N \| T95I |
| PV27654 | 2/7/21 | EPI_ISL_1708982 | B.1.429 | B.1.427/B.1.429 (20C.Cal) Detected. D614G Detected. P.2 Detected. | D614G \| E484K \| L452R \| S13I \| W152C |
| PV27671 | 2/7/21 | EPI_ISL_1708984 | B.1.526 | D614G Detected. Iota (B.1.526) Detected. | A701V \| D253G \| D614G \| E484K \| L5F \| T95I |
| PV27635 | 2/8/21 | EPI_ISL_1301431 | B.1.429 | B.1.427/B.1.429 (20C.Cal) Detected. D614G Detected. | D614G \| L452R \| S13I \| W152C |
| PV27643 | 2/8/21 | EPI_ISL_1300917 | B.1.526 | D614G Detected. Iota (B.1.526) Detected. | A701V \| D253G \| D614G \| E484K \| L5F \| T95I |
| PV27639 | 2/8/21 | EPI_ISL_1300806 | B.1.1.7 | Alpha (B.1.1.7) Detected. D614G Detected. | A570D \| A701V \| D253G \| D614G \| H69_V70del \| K1191N \| N501Y \| P681H \| S982A \| T716I \| T95I \| Y144del_Composite |
| PV27644 | 2/8/21 | EPI_ISL_1300918 | B.1.526 | D614G Detected. Iota (B.1.526) Detected. | A701V \| D253G \| D614G \| E484K \| L5F \| T95I |
| PV27645 | 2/8/21 | EPI_ISL_1300919 | B.1.526 | D614G Detected. Iota (B.1.526) Detected. | A701V \| D253G \| D614G \| E484K \| L5F \| T95I |
| PV27648 | 2/8/21 | EPI_ISL_1300920 | B.1.526 | D614G Detected. Iota (B.1.526) Detected. | A701V \| D253G \| D614G \| E484K \| L5F \| T95I |
| PV27646 | 2/8/21 | EPI_ISL_1300762 | B.1.526 | D614G Detected. Iota (B.1.526) Detected. | A701V \| D253G \| D614G \| E484K \| L5F \| T95I |
| PV27655 | 2/8/21 | EPI_ISL_1708985 | B.1.429 | B.1.427/B.1.429 (20C.Cal) Detected. D614G Detected. | D614G \| L452R \| S13I \| W152C |
| PV27653 | 2/9/21 | EPI_ISL_1301432 | B.1.429 | B.1.427/B.1.429 (20C.Cal) Detected. B.1.526.1 Detected. D614G Detected. | D614G \| D80G \| L452R \| S13I \| W152C |
| PV27659 | 2/9/21 | EPI_ISL_1300921 | B.1.526 | D614G Detected. Iota (B.1.526) Detected. | A701V \| D253G \| D614G \| E484K \| L5F \| T95I |
| PV27666 | 2/9/21 | EPI_ISL_1300811 | B.1.1.7 | Alpha (B.1.1.7) Detected. D614G Detected. | A570D \| D614G \| H69_V70del \| N501Y \| P681H \| S982A \| T716I \| Y144del_Composite |
| PV27683 | 2/9/21 | EPI_ISL_1708989 | B.1.526 | D614G Detected. Iota (B.1.526) Detected. | A701V \| D253G \| D614G \| E484K \| L5F \| T95I |
| PV27684 | 2/9/21 | EPI_ISL_1708991 | B.1.1.7 | Alpha (B.1.1.7) Detected. D614G Detected. | A570D \| D614G \| H69_V70del \| N501Y \| P681H \| S982A \| T716I \| Y144del_Composite |
| PV27685 | 2/9/21 | EPI_ISL_1708990 | B.1.526 | D614G Detected. Iota (B.1.526) Detected. | A701V \| D253G \| D614G \| E484K \| L5F \| T95I |
| PV27675 | 2/10/21 | EPI_ISL_5253420 | B.1.526 | D614G Detected. Iota (B.1.526) Detected. | A701V \| D253G \| D614G \| E484K \| L5F \| T95I |
| PV27676 | 2/10/21 | EPI_ISL_1300922 | B.1.526 | D614G Detected. Iota (B.1.526) Detected. | A701V \| D253G \| D614G \| L5F \| T95I |
| PV27682 | 2/10/21 | EPI_ISL_1300923 | B.1.526 | D614G Detected. Iota (B.1.526) Detected. | A701V \| D253G \| D614G \| E484K \| L5F \| T95I |
| PV27686 | 2/10/21 | EPI_ISL_1300924 | B.1.526 | D614G Detected. Iota (B.1.526) Detected. | A701V \| D253G \| D614G \| E484K \| L5F \| T95I |
| PV27680 | 2/10/21 | EPI_ISL_1301438 | B.1.429 | B.1.427/B.1.429 (20C.Cal) Detected. D614G Detected. | D614G \| L452R \| S13I \| W152C |
| PV27709 | 2/10/21 | EPI_ISL_1708996 | B.1.1.7 | Alpha (B.1.1.7) Detected. D614G Detected. | A570D \| D614G \| H69_V70del \| N501Y \| P681H \| S982A \| T716I |
| PV27691 | 2/11/21 | EPI_ISL_1300808 | B.1.1.7 | Alpha (B.1.1.7) Detected. D614G Detected. | A570D \| D614G \| H69_V70del \| N501Y \| P681H \| S982A \| T716I |
| PV27700 | 2/12/21 | EPI_ISL_1300849 | B.1.525 | D614G Detected. Eta (B.1.525) Detected. | D614G \| E484K \| H69_V70del \| Q677H \| Y144del_Composite |
| PV27705 | 2/13/21 | EPI_ISL_1300807 | B.1.1.7 | Alpha (B.1.1.7) Detected. D614G Detected. | A570D \| D614G \| H69_V70del \| N501Y \| P681H \| S982A \| T716I \| Y144del_Composite |
| PV28428 | 2/16/21 | EPI_ISL_1709006 | B.1.429 | B.1.427/B.1.429 (20C.Cal) Detected. D614G Detected. | D614G \| L452R \| S13I \| W152C |
| PV28444 | 2/17/21 | EPI_ISL_1709018 | B.1.1.7 | Alpha (B.1.1.7) Detected. D614G Detected. | A570D \| D614G \| H69_V70del \| K1191N \| N501Y \| P681H \| S982A \| T716I \| Y144del_Composite |
| PV28443 | 2/17/21 | EPI_ISL_1709014 | B.1.1.7 | Alpha (B.1.1.7) Detected. D614G Detected. | A570D \| D614G \| H69_V70del \| N501Y \| P681H \| S982A \| T716I \| Y144del_Composite |
| PV28450 | 2/17/21 | EPI_ISL_1709021 | B.1.1.7 | Alpha (B.1.1.7) Detected. D614G Detected. | A570D \| D614G \| H69_V70del \| N501Y \| P681H \| S982A \| T716I \| Y144del_Composite |
| PV28451 | 2/17/21 | EPI_ISL_1709027 | B.1.1.7 | Alpha (B.1.1.7) Detected. D614G Detected. | A570D \| D614G \| H69_V70del \| N501Y \| P681H \| S982A \| T716I \| Y144del_Composite |
| PV28459 | 2/17/21 | EPI_ISL_1709029 | B.1.1.7 | Alpha (B.1.1.7) Detected. D614G Detected. | A570D \| D614G \| H69_V70del \| L5F \| N501Y \| P681H \| S982A \| T716I \| Y144del_Composite |
| PV28467 | 2/18/21 | EPI_ISL_1709036 | B.1.429 | B.1.427/B.1.429 (20C.Cal) Detected. D614G Detected. | D614G \| L452R \| S13I \| W152C |
| PV28480 | 2/19/21 | EPI_ISL_1709043 | B.1.1.7 | Alpha (B.1.1.7) Detected. D614G Detected. | A570D \| D614G \| H69_V70del \| N501Y \| P681H \| S982A \| T716I \| Y144del_Composite |
| PV28503 | 2/22/21 | EPI_ISL_1709061 | B.1.427 | B.1.427/B.1.429 (20C.Cal) Detected. D614G Detected. | D614G \| L452R \| S13I \| W152C |
| PV28500 | 2/22/21 | EPI_ISL_1709060 | B.1.1.7 | Alpha (B.1.1.7) Detected. D614G Detected. | A570D \| D614G \| H69_V70del \| N501Y \| P681H \| S982A \| T716I \| Y144del_Composite |
| PV28508 | 2/22/21 | EPI_ISL_1709066 | B.1.1.7 | Alpha (B.1.1.7) Detected. D614G Detected. | A570D \| A701V \| D253G \| D614G \| H69_V70del \| N501Y \| P681H \| S982A \| T716I \| T95I |
| PV28496 | 2/22/21 | EPI_ISL_1709057 | B.1.2 | Broad USA Detected. D614G Detected. | D614G \| Q677P |
| PV28507 | 2/23/21 | EPI_ISL_5253379 | B.1.1.7 | Alpha (B.1.1.7) Detected. D614G Detected. P.2 Detected. | A570D \| D614G \| E484K \| H69_V70del \| L242_L244del \| N501Y \| P681H \| S982A \| T716I \| Y144del_Composite |
| PV28864 | 3/2/21 | EPI_ISL_1709159 | B.1.2 | Broad USA Detected. D614G Detected. | D614G \| H69_V70del_Dropout \| Q677P |
| PV35332 | 3/7/21 | EPI_ISL_5253382 | B.1.1.7 | Alpha (B.1.1.7) Detected. D614G Detected. Iota (B.1.526) Detected. P.2 Detected. | A570D \| A701V \| D253G \| D614G \| E484K \| H69_V70del \| N501Y \| P681H \| T716I \| T95I |
| PV35292 | 3/8/21 | EPI_ISL_5253401 | B.1.1.7 | Alpha (B.1.1.7) Detected. D614G Detected. | A570D \| D614G \| H69_V70del \| N501Y \| P681H \| S982A \| T716I \| Y144del_Composite |
| PV35293 | 3/8/21 | EPI_ISL_1709255 | C.37 | D614G Detected. | D614G \| T95I |
| PV35300 | 3/8/21 | EPI_ISL_1709254 | B.1.1.7 | Alpha (B.1.1.7) Detected. D614G Detected. | A570D \| D614G \| H69_V70del \| N501Y \| P681H \| S982A \| T716I \| Y144del_Composite |
| PV35339 | 3/8/21 | EPI_ISL_5253419 | B.1.429 | B.1.427/B.1.429 (20C.Cal) Detected. D614G Detected. | D614G \| L452R \| S13I \| W152C |
| PV35305 | 3/8/21 | EPI_ISL_1709262 | B.1.1.7 | Alpha (B.1.1.7) Detected. D614G Detected. | A570D \| D614G \| H69_V70del \| N501Y \| P681H \| S982A \| T716I \| Y144del_Composite |
| PV35319 | 3/9/21 | EPI_ISL_5253381 | B.1.1.7 | Alpha (B.1.1.7) Detected. D614G Detected. | A570D \| D614G \| H69_V70del \| N501Y \| P681H \| S982A \| T716I \| Y144del_Composite |
| PV35343 | 3/9/21 | EPI_ISL_1709285 | C.37 | D614G Detected. | D614G |
| PV35324 | 3/10/21 | EPI_ISL_1709286 | C.37 | D614G Detected. | D614G |
| PV35341 | 3/10/21 | EPI_ISL_1709289 | C.37 | D614G Detected. | D614G |
| PV35149 | 3/10/21 | EPI_ISL_1709296 | C.37 | D614G Detected. | D614G \| T95I |
| PV35328 | 3/10/21 | EPI_ISL_1709299 | B.1.1.7 | Alpha (B.1.1.7) Detected. D614G Detected. | A570D \| D614G \| H69_V70del \| N501Y \| P681H \| S982A \| T716I \| Y144del_Composite |
| PV35346 | 3/10/21 | EPI_ISL_1709297 | B.1.1.7 | Alpha (B.1.1.7) Detected. D614G Detected. | A570D \| D614G \| H69_V70del \| N501Y \| P681H \| S982A \| T716I \| Y144del_Composite |
| PV35350 | 3/11/21 | EPI_ISL_1709304 | B.1.1.7 | Alpha (B.1.1.7) Detected. D614G Detected. | A570D \| D614G \| H69_V70del \| N501Y \| P681H \| S982A \| T716I \| Y144del_Composite |
| PV35552 | 3/19/21 | EPI_ISL_1709529 | B.1.2 | Broad USA Detected. D614G Detected. | D614G \| E484-K/Q_Dropout \| H69_V70del_Dropout \| Q677P |
| PV35915 | 3/21/21 | EPI_ISL_1709556 | C.37 | D614G Detected. | D614G |
| PV35921 | 3/22/21 | EPI_ISL_5253270 | B.1.427 | B.1.427/B.1.429 (20C.Cal) Detected. D614G Detected. | D614G \| L452R \| S13I \| W152C |
| PV35926 | 3/22/21 | EPI_ISL_1709570 | B.1.1.7 | Alpha (B.1.1.7) Detected. D614G Detected. | A570D \| D614G \| H69_V70del \| N501Y \| P681H \| S982A \| T716I |
| PV35928 | 3/22/21 | EPI_ISL_1709572 | B.1.1.7 | Alpha (B.1.1.7) Detected. D614G Detected. | A570D \| D614G \| H69_V70del \| N501Y \| P681H \| S982A \| T716I \| Y144del_Composite |
| PV35929 | 3/22/21 | EPI_ISL_1709574 | B.1.1.7 | Alpha (B.1.1.7) Detected. D614G Detected. | A570D \| D614G \| H69_V70del \| N501Y \| P681H \| S982A \| T716I \| Y144del_Composite |
| PV35931 | 3/22/21 | EPI_ISL_1709577 | B.1.525 | D614G Detected. | D614G \| E484K \| H69_V70del_Dropout \| Q677H \| Y144del_Composite |
| PV35948 | 3/23/21 | EPI_ISL_1709627 | B.1.1.7 | Alpha (B.1.1.7) Detected. D614G Detected. | A570D \| D614G \| H69_V70del \| N501Y \| P681H \| S982A \| T716I \| Y144del_Composite |
| PV35953 | 3/23/21 | EPI_ISL_1709625 | P.1 | D614G Detected. Gamma (P.1) Detected. | D614G \| E484K \| K417T \| L18F \| N501Y |
| PV35955 | 3/23/21 | EPI_ISL_1709626 | P.1 | D614G Detected. Gamma (P.1) Detected. | D614G \| E484K \| K417T \| L18F \| N501Y |
| PV35947 | 3/24/21 | EPI_ISL_5253383 | B.1.1.7 | Alpha (B.1.1.7) Detected. D614G Detected. | A570D \| D614G \| H69_V70del \| N501Y \| P681H \| S982A \| T716I \| Y144del_Composite |
| PV35962 | 3/24/21 | EPI_ISL_1709654 | B.1.1.7 | Alpha (B.1.1.7) Detected. D614G Detected. | A570D \| D614G \| H69_V70del \| N501Y \| P681H \| S982A \| T716I \| Y144del_Composite |
| PV35963 | 3/24/21 | EPI_ISL_1709652 | B.1.1.7 | Alpha (B.1.1.7) Detected. D614G Detected. | A570D \| D614G \| H69_V70del \| N501Y \| P681H \| S982A \| T716I \| Y144del_Composite |
| PV35960 | 3/24/21 | EPI_ISL_1709650 | B.1.1.7 | Alpha (B.1.1.7) Detected. D614G Detected. | A570D \| D614G \| H69_V70del \| N501Y \| P681H \| S982A \| T716I \| Y144del_Composite |
| PV35957 | 3/25/21 | EPI_ISL_5253378 | B.1.1.7 | Alpha (B.1.1.7) Detected. D614G Detected. | A570D \| D614G \| H69_V70del \| L5F \| N501Y \| P681H \| S982A \| T716I \| Y144del_Composite |
| PV35958 | 3/25/21 | EPI_ISL_1709657 | B.1.1.7 | Alpha (B.1.1.7) Detected. D614G Detected. | A570D \| D614G \| H69_V70del \| N501Y \| P681H \| S982A \| T716I \| Y144del_Composite |
| PV35970 | 3/25/21 | EPI_ISL_5253272 | B.1.429 | B.1.427/B.1.429 (20C.Cal) Detected. D614G Detected. | D614G \| L452R \| S13I \| W152C |
| PV35966 | 3/25/21 | EPI_ISL_5253271 | B.1.427 | B.1.427/B.1.429 (20C.Cal) Detected. D614G Detected. | D614G \| L452R \| S13I \| W152C |
| PV35977 | 3/25/21 | EPI_ISL_1709686 | B.1.427 | B.1.427/B.1.429 (20C.Cal) Detected. D614G Detected. | D614G \| L452R \| S13I \| W152C |
| A259 | 3/29/21 | EPI_ISL_7307284 | P.1 | D614G Detected. Gamma (P.1) Detected. | D614G \| E484K \| K417T \| L18F \| N501Y |
| A260 | 3/29/21 | EPI_ISL_7307290 | P.1 | D614G Detected. Gamma (P.1) Detected. | D614G \| E484K \| K417T \| L18F \| N501Y |
| PV36288 | 3/29/21 | EPI_ISL_1709774 | B.1.351 | Beta (B.1.351) Detected. D614G Detected. | A701V \| D614G \| D80A \| E484K \| K417N \| L18F \| L242_L244del \| N501Y |
| PV36213 | 3/30/21 | EPI_ISL_1709785 | B.1.351 | Beta (B.1.351) Detected. Gamma (P.1) Detected. | A701V \| D80A \| E484K \| K417N \| K417T \| L18F \| N439K_Dropout \| N501Y |
| PV36885 | 4/4/21 | EPI_ISL_6492372 | B.1.525 | D614G Detected. | D614G \| E484K \| H69_V70del_Dropout \| Q677H |
| PV31898 | 4/19/21 | EPI_ISL_6492518 | B.1.525 | D614G Detected. | D614G \| E484K \| H69_V70del_Dropout \| Q677H \| Y144del_Composite |
| K12 | 4/23/21 | EPI_ISL_7476873 | C.37 | D614G Detected. | D614G |
| K2 | 4/23/21 | EPI_ISL_7476639 | C.37 | D614G Detected. | D614G |
| K3 | 4/23/21 | EPI_ISL_7476266 | C.37 | D614G Detected. | D614G |
| K5 | 4/23/21 | EPI_ISL_7476273 | C.37 | D614G Detected. | D614G |
| K6 | 4/23/21 | EPI_ISL_7476401 | C.37 | D614G Detected. | D614G |
| K7 | 4/23/21 | EPI_ISL_7476602 | C.37 | D614G Detected. | D614G |
| K18 | 4/26/21 | EPI_ISL_7476877 | C.37 | D614G Detected. | D614G |
| K19 | 4/26/21 | EPI_ISL_7476887 | C.37 | D614G Detected. | D614G |
| PV37801 | 4/29/21 | EPI_ISL_6492619 | B.1.525 | D614G Detected. | D614G \| E484K \| H69_V70del_Dropout \| Q677H |
| K13 | 5/1/21 | EPI_ISL_7476805 | C.37 | D614G Detected. | D614G |
| K22 | 5/4/21 | EPI_ISL_7476461 | C.37 | D614G Detected. | D614G \| T95I |
| K23 | 5/4/21 | EPI_ISL_7476811 | C.37 | D614G Detected. | D614G |
| K24 | 5/4/21 | EPI_ISL_7476815 | C.37 | D614G Detected. | D614G |
| PV31950 | 5/5/21 | EPI_ISL_6492667 | B.1.525 | D614G Detected. | D614G \| E484K \| H69_V70del_Dropout \| Q677H \| Y144del_Composite |
| PV31971 | 5/7/21 | EPI_ISL_6492691 | B.1.525 | D614G Detected. | D614G \| E484K \| H69_V70del_Dropout \| Q677H \| Y144del_Composite |
| K25 | 5/10/21 | EPI_ISL_7476823 | C.37 | D614G Detected. | D614G |
| K26 | 5/10/21 | EPI_ISL_7476833 | C.37 | D614G Detected. | D614G |
| K27 | 5/10/21 | EPI_ISL_7476836 | C.37 | D614G Detected. | D614G |
| H158 | 5/11/21 | EPI_ISL_7307276 | P.1 | D614G Detected. Gamma (P.1) Detected. | D614G \| E484K \| K417T \| L18F \| N501Y |
| K43 | 5/18/21 | EPI_ISL_7476423 | B.1.621 | D614G Detected. P.2 Detected. | D614G \| E484K \| N501Y \| P681H \| T95I \| Y144del_Composite |
| K47 | 5/18/21 | EPI_ISL_7476562 | B.1.621 | D614G Detected. P.2 Detected. | D614G \| E484K \| N501Y \| P681H \| T95I \| Y144del_Composite |
| K42 | 5/21/21 | EPI_ISL_7476627 | B.1.621 | D614G Detected. P.2 Detected. | D614G \| E484K \| N501Y \| P681H \| Q677H \| T95I \| Y144del_Composite |
| K45 | 5/21/21 | EPI_ISL_7476326 | B.1.621 | D614G Detected. P.2 Detected. | D614G \| E484K \| N501Y \| P681H \| T95I \| Y144del_Composite |
| K46 | 5/21/21 | EPI_ISL_7476345 | B.1.621 | D614G Detected. P.2 Detected. | D614G \| E484K \| N501Y \| P681H \| Q677H \| T95I \| Y144del_Composite |
| K49 | 5/29/21 | EPI_ISL_7476302 | B.1.621 | D614G Detected. P.2 Detected. | D614G \| E484K \| N501Y \| P681H \| T95I \| Y144del_Composite |
| K50 | 5/29/21 | EPI_ISL_7476680 | B.1.621 | D614G Detected. P.2 Detected. | D614G \| E484K \| N501Y \| P681H \| Q677H \| T95I \| Y144del_Composite |
| K53 | 5/29/21 | EPI_ISL_7476285 | B.1.621 | D614G Detected. P.2 Detected. | D614G \| E484K \| N501Y \| P681H \| T95I \| Y144del_Composite |
| K54 | 5/29/21 | EPI_ISL_7476482 | B.1.621 | D614G Detected. P.2 Detected. | D614G \| E484K \| N501Y \| P681H \| T95I \| Y144del_Composite |
| K56 | 5/29/21 | EPI_ISL_7476796 | B.1.621 | D614G Detected. P.2 Detected. | D614G \| E484K \| N501Y \| P681H \| T95I \| Y144del_Composite |
| K57 | 5/29/21 | EPI_ISL_7476365 | B.1.621 | D614G Detected. P.2 Detected. | D614G \| E484K \| N501Y \| P681H \| T95I \| Y144del_Composite |
| K68 | 6/2/21 | EPI_ISL_7476635 | P.1 | D614G Detected. Gamma (P.1) Detected. | D614G \| E484K \| K417T \| L18F \| N501Y |
| K59 | 6/4/21 | EPI_ISL_7476556 | P.1 | D614G Detected. Gamma (P.1) Detected. | D614G \| E484K \| K417T \| L18F \| N501Y |
| K61 | 6/4/21 | EPI_ISL_7476340 | B.1.621 | D614G Detected. P.2 Detected. | D614G \| E484K \| N501Y \| P681H \| T95I \| Y144del_Composite |
| K62 | 6/4/21 | EPI_ISL_7476181 | B.1.621 | D614G Detected. P.2 Detected. | D614G \| E484K \| N501Y \| P681H \| T95I \| Y144del_Composite |
| K63 | 6/4/21 | EPI_ISL_7476357 | B.1.621 | D614G Detected. P.2 Detected. | D614G \| E484K \| N501Y \| P681H \| T95I \| Y144del_Composite |
| H100 | 6/16/21 | EPI_ISL_7307242 | P.1 | D614G Detected. Gamma (P.1) Detected. | D614G \| E484K \| K417T \| L18F \| N501Y |
| H99 | 6/16/21 | EPI_ISL_7307289 | P.1 | D614G Detected. Gamma (P.1) Detected. | D614G \| E484K \| K417T \| L18F \| N501Y |
| K95 | 6/21/21 | EPI_ISL_7476174 | P.1 | D614G Detected. Gamma (P.1) Detected. | D614G \| E484K \| K417T \| L18F \| N501Y |
| K92 | 6/23/21 | EPI_ISL_7476661 | P.1 | D614G Detected. Gamma (P.1) Detected. | D614G \| E484K \| K417T \| L18F \| N501Y |
| H119 | 7/4/21 | EPI_ISL_7307241 | P.1 | D614G Detected. Gamma (P.1) Detected. | D614G \| E484K \| K417T \| L18F \| N501Y |
| K115 | 7/6/21 | EPI_ISL_7476570 | P.1 | D614G Detected. Gamma (P.1) Detected. | D614G \| E484K \| K417T \| L18F \| N501Y |
| V17 | 7/22/21 | EPI_ISL_12084490 | P.1 | D614G Detected. Gamma (P.1) Detected. | D614G \| E484K \| K417T \| L18F \| N501Y |
| K38 | 7/28/21 | EPI_ISL_7476253 | B.1.621 | D614G Detected. P.2 Detected. | D614G \| E484K \| N501Y \| P681H \| T95I \| Y144del_Composite |
| PV40356 | 11/1/21 | EPI_ISL_6494430 | AY.44 | D614G Detected. Delta (B.1.617.2) Detected. | D614G \| L452R \| P681R \| T19R \| T478K |
| PV40371 | 11/1/21 | EPI_ISL_6494434 | AY.103 | D614G Detected. Delta (B.1.617.2) Detected. | D614G \| L452R \| P681R \| T19R \| T478K |
| PV40360 | 11/1/21 | EPI_ISL_6494432 | AY.103 | D614G Detected. Delta (B.1.617.2) Detected. | D614G \| L452R \| P681R \| T19R \| T478K |
| PV40358 | 11/1/21 | EPI_ISL_6494431 | AY.118 | D614G Detected. Delta (B.1.617.2) Detected. | D614G \| L452R \| P681R \| T19R \| T478K \| T95I |
| PV40364 | 11/1/21 | EPI_ISL_6494433 | AY.25 | D614G Detected. Delta (B.1.617.2) Detected. | D614G \| L452R \| P681R \| T19R \| T478K |
| PV40351 | 11/2/21 | EPI_ISL_12131519 | B.1.617.2 | D614G Detected. Delta (B.1.617.2) Detected. | D614G \| L452R \| P681R \| T19R \| T478K |
| PV40365 | 11/2/21 | EPI_ISL_6494439 | AY.103 | D614G Detected. Delta (B.1.617.2) Detected. | D614G \| L452R \| P681R \| T19R \| T478K |
| PV40368 | 11/2/21 | EPI_ISL_6494440 | AY.103 | D614G Detected. Delta (B.1.617.2) Detected. | D614G \| L452R \| P681R \| T19R \| T478K \| T95I |
| PV40344 | 11/2/21 | EPI_ISL_6494437 | AY.100 | D614G Detected. Delta (B.1.617.2) Detected. | D614G \| L452R \| P681R \| T19R \| T478K \| T95I |
| PV40345 | 11/2/21 | EPI_ISL_6494438 | AY.103 | D614G Detected. Delta (B.1.617.2) Detected. | D614G \| L452R \| P681R \| T19R \| T478K |
| PV40331 | 11/2/21 | EPI_ISL_6494436 | AY.100 | D614G Detected. Delta (B.1.617.2) Detected. | D614G \| L452R \| P681R \| T19R \| T478K \| T95I |
| PV40333 | 11/3/21 | Mixed | AY.103 | D614G Detected. Delta (B.1.617.2) Detected. | D614G \| L452R \| P681R \| T19R \| T478K |
| PV40337 | 11/3/21 | Mixed | AY.103 | D614G Detected. Delta (B.1.617.2) Detected. | D614G \| L452R \| P681R \| T19R \| T478K \| T95I |
| PV40330 | 11/3/21 | Mixed | B.1.617.2 | D614G Detected. Delta (B.1.617.2) Detected. | D614G \| L452R \| P681R \| T19R \| T478K \| T95I |
| PV40328 | 11/3/21 | EPI_ISL_6494447 | AY.44 | D614G Detected. Delta (B.1.617.2) Detected. | D614G \| L452R \| P681R \| T19R \| T478K |
| PV40370 | 11/4/21 | EPI_ISL_6494450 | AY.103 | D614G Detected. Delta (B.1.617.2) Detected. | D614G \| L452R \| P681R \| T19R \| T478K |
| PV40336 | 11/4/21 | Mixed | AY.121 | D614G Detected. Delta (B.1.617.2) Detected. | D614G \| L452R \| P681R \| T19R \| T478K \| T95I |
| PV40334 | 11/4/21 | EPI_ISL_6494449 | AY.121 | D614G Detected. Delta (B.1.617.2) Detected. | D614G \| L452R \| P681R \| T19R \| T478K \| T95I |
| PV40329 | 11/4/21 | EPI_ISL_6494448 | AY.103 | D614G Detected. Delta (B.1.617.2) Detected. | D614G \| L452R \| P681R \| T19R \| T478K |
| PV40327 | 11/4/21 | EPI_ISL_12131518 | AY.3 | D614G Detected. Delta (B.1.617.2) Detected. | D614G \| L452R \| P681R \| T19R \| T478K |
| PV40338 | 11/5/21 | EPI_ISL_6494454 | AY.119 | D614G Detected. Delta (B.1.617.2) Detected. | D614G \| L452R \| P681R \| T19R \| T478K |
| PV40317 | 11/5/21 | EPI_ISL_6494451 | B.1.617.2 | D614G Detected. Delta (B.1.617.2) Detected. | D614G \| L452R \| P681R \| T19R \| T478K \| T95I |
| PV40325 | 11/5/21 | EPI_ISL_6494453 | AY.117 | D614G Detected. | D614G \| E484-K/Q_Dropout \| H69_V70del_Dropout \| L452R \| T478K \| T95I |
| PV40341 | 11/5/21 | EPI_ISL_6494455 | AY.33 | D614G Detected. Delta (B.1.617.2) Detected. | D614G \| L452R \| P681R \| T19R \| T478K |
| PV40319 | 11/5/21 | EPI_ISL_6494452 | AY.33 | D614G Detected. Delta (B.1.617.2) Detected. | D614G \| L452R \| P681R \| T19R \| T478K |
| PV40323 | 11/5/21 | EPI_ISL_12131516 | AY.44 | D614G Detected. Delta (B.1.617.2) Detected. | D614G \| L452R \| P681R \| T19R \| T478K |
| PV41850 | 11/6/21 | EPI_ISL_7907853 | AY.103 | D614G Detected. Delta (B.1.617.2) Detected. | D614G \| L452R \| T19R \| T478K |
| PV41851 | 11/6/21 | EPI_ISL_7907956 | AY.122 | D614G Detected. Delta (B.1.617.2) Detected. | D614G \| L452R \| P681R \| T19R \| T478K \| T95I |
| PV40315 | 11/6/21 | EPI_ISL_6494458 | AY.39 | D614G Detected. Delta (B.1.617.2) Detected. | D614G \| L452R \| P681R \| T19R \| T478K \| T95I |
| PV40324 | 11/7/21 | EPI_ISL_12131517 | AY.103 | D614G Detected. Delta (B.1.617.2) Detected. | D614G \| L452R \| P681R \| T19R \| T478K |
| PV40318 | 11/7/21 | EPI_ISL_12131515 | AY.103 | D614G Detected. Delta (B.1.617.2) Detected. | D614G \| L452R \| P681R \| T19R \| T478K |
| PV41854 | 11/7/21 | EPI_ISL_7907957 | AY.103 | D614G Detected. Delta (B.1.617.2) Detected. | D614G \| L452R \| N439K_Dropout \| P681R \| T19R \| T478K \| T95I |
| PV41853 | 11/8/21 | EPI_ISL_7907923 | AY.119.2 | D614G Detected. Delta (B.1.617.2) Detected. | D614G \| L452R \| P681R \| T19R \| T478K \| T95I |
| PV41857 | 11/8/21 | EPI_ISL_7907902 | AY.3 | D614G Detected. Delta (B.1.617.2) Detected. | D614G \| L452R \| P681R \| T19R \| T478K |
| PV41860 | 11/8/21 | EPI_ISL_7907852 | AY.103 | D614G Detected. Delta (B.1.617.2) Detected. | D614G \| L452R \| P681R \| T19R \| T478K |
| PV41870 | 11/9/21 | EPI_ISL_7907960 | AY.47 | D614G Detected. Delta (B.1.617.2) Detected. | D614G \| L452R \| P681R \| T19R \| T478K |
| PV41889 | 11/9/21 | Mixed | AY.43 | D614G Detected. Delta (B.1.617.2) Detected. | D614G \| L452R \| P681R \| T19R \| T478K |
| PV41900 | 11/9/21 | EPI_ISL_7907929 | AY.109 | D614G Detected. Delta (B.1.617.2) Detected. | D614G \| L452R \| P681R \| T19R \| T478K \| T95I |
| PV41868 | 11/9/21 | EPI_ISL_7907958 | AY.20 | D614G Detected. Delta (B.1.617.2) Detected. | D614G \| L452R \| P681R \| T19R \| T478K \| T95I |
| PV41881 | 11/9/21 | EPI_ISL_7907892 | AY.25 | D614G Detected. Delta (B.1.617.2) Detected. | D614G \| L452R \| L5F \| P681R \| T19R \| T478K |
| PV41871 | 11/9/21 | EPI_ISL_7907914 | AY.25.1 | D614G Detected. Delta (B.1.617.2) Detected. | D614G \| L452R \| P681R \| T19R \| T478K |
| PV41903 | 11/10/21 | EPI_ISL_7907965 | AY.26 | D614G Detected. Delta (B.1.617.2) Detected. | D614G \| L452R \| P681R \| T19R \| T478K |
| PV41904 | 11/10/21 | EPI_ISL_7907854 | AY.26 | D614G Detected. Delta (B.1.617.2) Detected. | D614G \| L452R \| P681R \| T19R \| T478K |
| PV41872 | 11/10/21 | EPI_ISL_7907883 | AY.47 | D614G Detected. Delta (B.1.617.2) Detected. | D614G \| L452R \| P681R \| T19R \| T478K |
| PV41878 | 11/10/21 | EPI_ISL_7907884 | AY.47 | D614G Detected. Delta (B.1.617.2) Detected. | D614G \| L452R \| P681R \| T19R \| T478K |
| PV41869 | 11/10/21 | EPI_ISL_7907882 | AY.47 | D614G Detected. | D614G \| E484-K/Q_Dropout \| H69_V70del_Dropout \| L452R \| T478K |
| PV41895 | 11/10/21 | EPI_ISL_7907926 | AY.118 | D614G Detected. Delta (B.1.617.2) Detected. | D614G \| L452R \| P681R \| T19R \| T478K \| T95I |
| PV41874 | 11/10/21 | EPI_ISL_7907924 | AY.103 | D614G Detected. Delta (B.1.617.2) Detected. | D614G \| L452R \| P681R \| T19R \| T478K |
| PV41898 | 11/10/21 | EPI_ISL_7907927 | AY.103 | D614G Detected. Delta (B.1.617.2) Detected. | D614G \| L452R \| P681R \| T19R \| T478K |
| PV41896 | 11/10/21 | EPI_ISL_7908115 | AY.4 | D614G Detected. Delta (B.1.617.2) Detected. | D614G \| L452R \| P681R \| T19R \| T478K \| T95I |
| PV41862 | 11/10/21 | EPI_ISL_7907988 | AY.44 | D614G Detected. Delta (B.1.617.2) Detected. | D614G \| H69_V70del \| L452R \| P681R \| T19R \| T478K |
| PV41861 | 11/10/21 | EPI_ISL_7907982 | AY.39 | D614G Detected. Delta (B.1.617.2) Detected. | D614G \| L452R \| P681R \| T19R \| T478K \| T95I |
| PV41876 | 11/10/21 | EPI_ISL_7907915 | AY.103 | D614G Detected. Delta (B.1.617.2) Detected. | D614G \| L452R \| P681R \| T19R \| T478K |
| PV41899 | 11/11/21 | EPI_ISL_7907928 | AY.100 | D614G Detected. Delta (B.1.617.2) Detected. | D614G \| L452R \| P681R \| T19R \| T478K \| T95I |
| PV41928 | 11/11/21 | EPI_ISL_7907886 | AY.103 | D614G Detected. Delta (B.1.617.2) Detected. | D614G \| L452R \| P681R \| T19R \| T478K |
| PV41887 | 11/11/21 | EPI_ISL_7907962 | AY.44 | D614G Detected. Delta (B.1.617.2) Detected. | D614G \| L452R \| P681R \| T19R \| T478K |
| PV41893 | 11/11/21 | EPI_ISL_7907963 | AY.103 | D614G Detected. Delta (B.1.617.2) Detected. | D614G \| L452R \| P681R \| T19R \| T478K \| T95I |
| PV41918 | 11/11/21 | EPI_ISL_7907966 | AY.25.1 | D614G Detected. Delta (B.1.617.2) Detected. | D614G \| L452R \| P681R \| T19R \| T478K |
| PV41912 | 11/11/21 | EPI_ISL_7907931 | AY.3.3 | D614G Detected. Delta (B.1.617.2) Detected. | D614G \| L452R \| P681R \| T19R \| T478K \| T95I |
| PV41886 | 11/11/21 | EPI_ISL_7907961 | AY.3 | D614G Detected. Delta (B.1.617.2) Detected. | D614G \| L452R \| P681R \| T19R \| T478K |
| PV41902 | 11/11/21 | EPI_ISL_7907964 | AY.3 | D614G Detected. | D614G \| E484-K/Q_Dropout \| H69_V70del_Dropout \| L452R \| T478K |
| PV41873 | 11/11/21 | EPI_ISL_7908114 | B.1.351 | D614G Detected. | A701V \| D614G \| E484K \| H69_V70del_Dropout \| N439K_Dropout \| N501Y |
| PV41925 | 11/12/21 | EPI_ISL_7907903 | AY.9.2 | D614G Detected. Delta (B.1.617.2) Detected. | D614G \| L452R \| P681R \| T19R \| T478K |
| PV41890 | 11/12/21 | EPI_ISL_7907925 | AY.126 | D614G Detected. Delta (B.1.617.2) Detected. | D614G \| L452R \| P681R \| T19R \| T478K \| T95I |
| PV41908 | 11/12/21 | EPI_ISL_7907916 | AY.103 | D614G Detected. Delta (B.1.617.2) Detected. | D614G \| L452R \| P681R \| T19R \| T478K \| T95I |
| PV41917 | 11/12/21 | EPI_ISL_7907917 | AY.103 | D614G Detected. Delta (B.1.617.2) Detected. | D614G \| L452R \| P681R \| T19R \| T478K |
| PV41923 | 11/12/21 | Mixed | AY.103 | D614G Detected. Delta (B.1.617.2) Detected. | D614G \| L452R \| P681R \| T19R \| T478K |
| PV41926 | 11/12/21 | EPI_ISL_7907967 | AY.3 | D614G Detected. Delta (B.1.617.2) Detected. | D614G \| L452R \| P681R \| T19R \| T478K |
| PV41929 | 11/13/21 | EPI_ISL_7907932 | AY.103 | D614G Detected. Delta (B.1.617.2) Detected. | D614G \| L452R \| P681R \| T19R \| T478K |
| PV42315 | 11/13/21 | EPI_ISL_7907969 | AY.103 | D614G Detected. Delta (B.1.617.2) Detected. | D614G \| L452R \| P681R \| T19R \| T478K |
| PV42312 | 11/13/21 | EPI_ISL_7907936 | AY.103 | D614G Detected. Delta (B.1.617.2) Detected. | D614G \| L452R \| P681R \| T19R \| T478K |
| PV42295 | 11/14/21 | EPI_ISL_7907887 | AY.103 | D614G Detected. Delta (B.1.617.2) Detected. | D614G \| L452R \| P681R \| T19R \| T478K |
| PV42311 | 11/14/21 | EPI_ISL_7907893 | AY.44 | D614G Detected. Delta (B.1.617.2) Detected. | D614G \| L452R \| P681R \| T19R \| T478K |
| PV42313 | 11/15/21 | EPI_ISL_7907968 | AY.119 | D614G Detected. | D614G \| E484-K/Q_Dropout \| H69_V70del_Dropout \| L452R \| T478K \| T95I |
| PV42299 | 11/15/21 | EPI_ISL_7907904 | AY.25 | D614G Detected. Delta (B.1.617.2) Detected. | D614G \| L452R \| P681R \| T19R \| T478K |
| PV42300 | 11/15/21 | EPI_ISL_7907905 | AY.106 | D614G Detected. Delta (B.1.617.2) Detected. | D614G \| L452R \| L5F \| P681R \| T19R \| T478K |
| PV42317 | 11/16/21 | EPI_ISL_7907937 | AY.44 | D614G Detected. Delta (B.1.617.2) Detected. | D614G \| L452R \| P681R \| T19R \| T478K |
| PV42341 | 11/16/21 | EPI_ISL_7907895 | AY.122 | D614G Detected. Delta (B.1.617.2) Detected. | D614G \| L452R \| P681R \| T19R \| T478K |
| PV42351 | 11/17/21 | Mixed | AY.75 | D614G Detected. Delta (B.1.617.2) Detected. | D614G \| L452R \| N439K_Dropout \| P681R \| T19R \| T478K |
| PV42345 | 11/17/21 | Mixed | AY.39 | D614G Detected. Delta (B.1.617.2) Detected. | D614G \| L452R \| P681R \| T19R \| T478K \| T95I |
| PV42355 | 11/17/21 | EPI_ISL_7907920 | AY.25 | D614G Detected. Delta (B.1.617.2) Detected. | D614G \| L452R \| P681R \| T19R \| T478K |
| PV42348 | 11/17/21 | EPI_ISL_7907888 | AY.43 | D614G Detected. Delta (B.1.617.2) Detected. | D614G \| L452R \| P681R \| T19R \| T478K \| T95I |
| PV42346 | 11/17/21 | EPI_ISL_7907987 | AY.44 | D614G Detected. Delta (B.1.617.2) Detected. | D614G \| L452R \| P681R \| T19R \| T478K |
| PV42340 | 11/17/21 | EPI_ISL_7907940 | AY.118 | D614G Detected. Delta (B.1.617.2) Detected. | D614G \| L452R \| P681R \| T19R \| T478K \| T95I |
| PV42335 | 11/17/21 | EPI_ISL_7907919 | AY.25 | D614G Detected. Delta (B.1.617.2) Detected. | D614G \| L452R \| P681R \| T19R \| T478K |
| PV42336 | 11/17/21 | EPI_ISL_7907970 | AY.3.1 | D614G Detected. Delta (B.1.617.2) Detected. | D614G \| L452R \| P681R \| T19R \| T478K |
| PV42338 | 11/17/21 | EPI_ISL_7907894 | AY.3.1 | D614G Detected. Delta (B.1.617.2) Detected. | D614G \| L452R \| P681R \| T19R \| T478K |
| PV42356 | 11/17/21 | Mixed | AY.43 | D614G Detected. Delta (B.1.617.2) Detected. | D614G \| L452R \| P681R \| T19R \| T478K |
| PV42337 | 11/17/21 | EPI_ISL_7907939 | AY.122 | D614G Detected. Delta (B.1.617.2) Detected. | D614G \| L452R \| P681R \| T19R \| T478K |
| PV42360 | 11/18/21 | EPI_ISL_7908121 | AY.103 | D614G Detected. Delta (B.1.617.2) Detected. | D614G \| L452R \| P681R \| T19R \| T478K |
| PV42362 | 11/18/21 | EPI_ISL_7908129 | AY.103 | D614G Detected. Delta (B.1.617.2) Detected. | D614G \| L452R \| P681R \| T19R \| T478K |
| PV42364 | 11/18/21 | EPI_ISL_7907971 | AY.100 | D614G Detected. Delta (B.1.617.2) Detected. | D614G \| N439K_Dropout \| P681R \| T19R \| T478K \| T95I |
| PV42366 | 11/18/21 | EPI_ISL_7908131 | AY.103 | D614G Detected. Delta (B.1.617.2) Detected. | D614G \| L452R \| P681R \| T19R \| T478K |
| PV42378 | 11/19/21 | EPI_ISL_7907941 | AY.103 | D614G Detected. Delta (B.1.617.2) Detected. | D614G \| L452R \| T19R \| T478K |
| PV42382 | 11/19/21 | EPI_ISL_7908120 | AY.3 | D614G Detected. Delta (B.1.617.2) Detected. | D614G \| L452R \| P681R \| T19R \| T478K |
| PV42375 | 11/19/21 | EPI_ISL_7907972 | AY.3 | D614G Detected. Delta (B.1.617.2) Detected. | D614G \| L452R \| P681R \| T19R \| T478K |
| PV42377 | 11/19/21 | EPI_ISL_7907889 | AY.25.1 | D614G Detected. Delta (B.1.617.2) Detected. | D614G \| L452R \| P681R \| T19R \| T478K |
| PV45513 | 12/13/21 | EPI_ISL_7908098 | BA.1 | D614G Detected. Omicron (B.1.1.529) Detected. | D614G \| H69_V70del_Dropout \| K417N \| N439K_Dropout \| P681H \| S477N \| T478K \| T95I \| Y144del_Composite |
| PV45522 | 12/13/21 | EPI_ISL_7908067 | BA.1 | D614G Detected. Omicron (B.1.1.529) Detected. | D614G \| H69_V70del_Dropout \| K417N \| N439K_Dropout \| P681H \| S477N \| T478K \| T95I \| Y144del_Composite |
| PV45515 | 12/13/21 | EPI_ISL_7908066 | BA.1 | D614G Detected. Omicron (B.1.1.529) Detected. | D614G \| H69_V70del_Dropout \| K417N \| N439K_Dropout \| P681H \| S477N \| T478K \| T95I \| Y144del_Composite |
| PV45462 | 12/13/21 | EPI_ISL_7908017 | BA.1 | D614G Detected. Omicron (B.1.1.529) Detected. | D614G \| H69_V70del_Dropout \| K417N \| N439K_Dropout \| P681H \| S477N \| T478K \| T95I \| Y144del_Composite |
| PV45475 | 12/14/21 | EPI_ISL_7907891 | AY.25.1 | D614G Detected. Delta (B.1.617.2) Detected. | D614G \| L452R \| P681R \| T19R \| T478K |
| PV45464 | 12/14/21 | EPI_ISL_7907980 | AY.100 | D614G Detected. Delta (B.1.617.2) Detected. | D614G \| L452R \| P681R \| T19R \| T478K \| T95I |
| PV45451 | 12/14/21 | EPI_ISL_7907986 | AY.44 | D614G Detected. Delta (B.1.617.2) Detected. | D614G \| E484-K/Q_Dropout \| L452R \| P681R \| T19R |
| PV45468 | 12/14/21 | EPI_ISL_7907954 | AY.43 | D614G Detected. Delta (B.1.617.2) Detected. | D614G \| L452R \| P681R \| T19R \| T478K |
| PV45473 | 12/14/21 | EPI_ISL_7907955 | AY.3 | D614G Detected. Delta (B.1.617.2) Detected. | D614G \| L452R \| P681R \| T19R \| T478K |
| PV45430 | 12/14/21 | EPI_ISL_7907952 | AY.43 | D614G Detected. Delta (B.1.617.2) Detected. | D614G \| L452R \| P681R \| T19R \| T478K |
| PV45503 | 12/14/21 | EPI_ISL_7908060 | AY.3 | D614G Detected. Delta (B.1.617.2) Detected. | D614G \| L452R \| P681R \| T19R \| T478K |
| PV45448 | 12/14/21 | EPI_ISL_7907953 | AY.103 | D614G Detected. Delta (B.1.617.2) Detected. | D614G \| L452R \| P681R \| T19R \| T478K |
| PV45453 | 12/14/21 | EPI_ISL_7908003 | BA.1.1 | D614G Detected. Omicron (B.1.1.529) Detected. | D614G \| H69_V70del_Dropout \| K417N \| N439K_Dropout \| P681H \| S477N \| T478K \| T95I \| Y144del_Composite |
| PV45497 | 12/14/21 | EPI_ISL_7908074 | BA.1 | D614G Detected. Omicron (B.1.1.529) Detected. | D614G \| H69_V70del_Dropout \| K417N \| N439K_Dropout \| P681H \| S477N \| T478K \| T95I \| Y144del_Composite |
| PV45477 | 12/14/21 | EPI_ISL_7908090 | BA.1 | D614G Detected. Omicron (B.1.1.529) Detected. | D614G \| H69_V70del_Dropout \| K417N \| N439K_Dropout \| P681H \| S477N \| T478K \| T95I \| Y144del_Composite |
| PV45466 | 12/14/21 | EPI_ISL_7908088 | BA.1.1 | D614G Detected. Omicron (B.1.1.529) Detected. | D614G \| H69_V70del_Dropout \| K417N \| N439K_Dropout \| P681H \| S477N \| T478K \| T95I \| Y144del_Composite |
| PV45454 | 12/14/21 | EPI_ISL_11178969 | BA.1.1 | D614G Detected. Delta (AY.1/AY.2) Detected. | D614G \| K417N \| P681H \| S477N \| T19R \| T478K \| T95I |
| PV45460 | 12/14/21 | EPI_ISL_7908004 | BA.1 | D614G Detected. Omicron (B.1.1.529) Detected. | D614G \| H69_V70del_Dropout \| K417N \| N439K_Dropout \| P681H \| S477N \| T478K \| T95I \| Y144del_Composite |
| PV45452 | 12/14/21 | EPI_ISL_7908043 | BA.1 | D614G Detected. Omicron (B.1.1.529) Detected. | D614G \| H69_V70del_Dropout \| K417N \| N439K_Dropout \| P681H \| S477N \| T478K \| T95I \| Y144del_Composite |
| PV45446 | 12/14/21 | EPI_ISL_7908065 | BA.1 | D614G Detected. Omicron (B.1.1.529) Detected. | D614G \| H69_V70del_Dropout \| K417N \| N439K_Dropout \| P681H \| S477N \| T478K \| T95I \| Y144del_Composite |
| PV45489 | 12/14/21 | EPI_ISL_7908096 | BA.1.1 | D614G Detected. Omicron (B.1.1.529) Detected. | D614G \| H69_V70del_Dropout \| K417N \| N439K_Dropout \| P681H \| S477N \| T478K \| T95I \| Y144del_Composite |
| PV45467 | 12/14/21 | EPI_ISL_7908045 | BA.1 | D614G Detected. Omicron (B.1.1.529) Detected. | D614G \| H69_V70del \| K417N \| N439K_Dropout \| P681H \| S477N \| T478K \| T95I \| Y144del_Composite |
| PV45459 | 12/14/21 | EPI_ISL_7908070 | BA.1 | D614G Detected. Omicron (B.1.1.529) Detected. | D614G \| H69_V70del_Dropout \| K417N \| N439K_Dropout \| P681H \| S477N \| T478K \| T95I \| Y144del_Composite |
| PV45471 | 12/14/21 | EPI_ISL_7908089 | BA.1 | D614G Detected. | D614G \| E484-K/Q_Dropout \| H69_V70del_Dropout \| N439K_Dropout \| S477N \| T478K \| T95I |
| PV45472 | 12/14/21 | EPI_ISL_7908071 | BA.1 | D614G Detected. Omicron (B.1.1.529) Detected. | A701V \| D614G \| H69_V70del \| K417N \| N439K_Dropout \| P681H \| S477N \| T478K \| T95I \| Y144del_Composite |
| PV45458 | 12/14/21 | EPI_ISL_7908086 | BA.1.1 | D614G Detected. Omicron (B.1.1.529) Detected. | D614G \| H69_V70del_Dropout \| K417N \| N439K_Dropout \| P681H \| S477N \| T478K \| T95I \| Y144del_Composite |
| PV45465 | 12/14/21 | EPI_ISL_7908087 | BA.1 | D614G Detected. Omicron (B.1.1.529) Detected. | D614G \| H69_V70del_Dropout \| K417N \| N439K_Dropout \| P681H \| S477N \| T478K \| T95I \| Y144del_Composite |
| PV45478 | 12/14/21 | EPI_ISL_7908091 | BA.1 | D614G Detected. Omicron (B.1.1.529) Detected. | D614G \| H69_V70del_Dropout \| K417N \| N439K_Dropout \| P681H \| S477N \| T478K \| T95I \| Y144del_Composite |
| PV45506 | 12/14/21 | EPI_ISL_7908109 | BA.1 | D614G Detected. Omicron (B.1.1.529) Detected. | D614G \| H69_V70del_Dropout \| K417N \| N439K_Dropout \| P681H \| S477N \| T478K \| T95I \| Y144del_Composite |
| PV45490 | 12/14/21 | EPI_ISL_7908097 | BA.1 | D614G Detected. Omicron (B.1.1.529) Detected. | D614G \| H69_V70del_Dropout \| K417N \| N439K_Dropout \| P681H \| S477N \| T478K \| T95I \| Y144del_Composite |
| PV45482 | 12/14/21 | EPI_ISL_7908094 | BA.1 | D614G Detected. Omicron (B.1.1.529) Detected. | A701V \| D614G \| E484-K/Q_Dropout \| H69_V70del_Dropout \| K417N \| N439K_Dropout \| P681H \| S477N \| T478K \| T95I |
| PV45433 | 12/14/21 | EPI_ISL_7908016 | BA.1 | D614G Detected. Omicron (B.1.1.529) Detected. | A701V \| D614G \| H69_V70del_Dropout \| K417N \| N439K_Dropout \| P681H \| S477N \| T478K \| T95I \| Y144del_Composite |
| PV45432 | 12/14/21 | EPI_ISL_7908072 | BA.1.1 | D614G Detected. Omicron (B.1.1.529) Detected. | D614G \| H69_V70del_Dropout \| K417N \| N439K_Dropout \| P681H \| S477N \| T478K \| T95I \| Y144del_Composite |
| PV45484 | 12/14/21 | EPI_ISL_7908047 | BA.1.1 | D614G Detected. Omicron (B.1.1.529) Detected. | D614G \| H69_V70del_Dropout \| K417N \| N439K_Dropout \| P681H \| S477N \| T478K \| T95I \| Y144del_Composite |
| PV45438 | 12/14/21 | EPI_ISL_7908085 | BA.1 | D614G Detected. Omicron (B.1.1.529) Detected. | D614G \| H69_V70del_Dropout \| K417N \| N439K_Dropout \| P681H \| S477N \| T478K \| T95I \| Y144del_Composite |
| PV45479 | 12/14/21 | EPI_ISL_7908046 | BA.1 | D614G Detected. Omicron (B.1.1.529) Detected. | D614G \| H69_V70del_Dropout \| K417N \| N439K_Dropout \| P681H \| S477N \| T478K \| T95I \| Y144del_Composite |
| PV45463 | 12/14/21 | EPI_ISL_7908044 | BA.1 | D614G Detected. Omicron (B.1.1.529) Detected. | D614G \| H69_V70del_Dropout \| K417N \| N439K_Dropout \| P681H \| S477N \| T478K \| T95I \| Y144del_Composite |
| PV45510 | 12/14/21 | EPI_ISL_7908049 | BA.1 | D614G Detected. | D614G \| E484-K/Q_Dropout \| H69_V70del_Dropout \| N439K_Dropout \| S477N \| T478K \| T95I |
| PV45514 | 12/14/21 | EPI_ISL_7908050 | BA.1.1 | D614G Detected. Omicron (B.1.1.529) Detected. | D614G \| H69_V70del_Dropout \| K417N \| P681H \| S477N \| T478K \| T95I \| Y144del_Composite |
| PV45440 | 12/14/21 | EPI_ISL_7908002 | BA.1 | D614G Detected. Omicron (B.1.1.529) Detected. | D614G \| H69_V70del_Dropout \| K417N \| N439K_Dropout \| P681H \| S477N \| T478K \| T95I \| Y144del_Composite |
| PV45505 | 12/14/21 | EPI_ISL_7908063 | BA.1 | D614G Detected. Omicron (B.1.1.529) Detected. | D614G \| H69_V70del_Dropout \| K417N \| N439K_Dropout \| P681H \| S477N \| T478K \| T95I \| Y144del_Composite |
| PV45500 | 12/14/21 | EPI_ISL_7908075 | BA.1 | D614G Detected. Omicron (B.1.1.529) Detected. | D614G \| H69_V70del \| K417N \| N439K_Dropout \| P681H \| S477N \| T478K \| T95I \| Y144del_Composite |
| PV45480 | 12/14/21 | EPI_ISL_7908092 | BA.1 | D614G Detected. Omicron (B.1.1.529) Detected. | D614G \| H69_V70del_Dropout \| K417N \| N439K_Dropout \| P681H \| S477N \| T478K \| T95I \| Y144del_Composite |
| PV45449 | 12/14/21 | EPI_ISL_7908073 | BA.1 | D614G Detected. Omicron (B.1.1.529) Detected. | D614G \| H69_V70del_Dropout \| K417N \| N439K_Dropout \| P681H \| S477N \| T478K \| T95I \| Y144del_Composite |
| PV45492 | 12/14/21 | EPI_ISL_7908048 | BA.1 | D614G Detected. Omicron (B.1.1.529) Detected. | A701V \| D614G \| H69_V70del_Dropout \| K417N \| N439K_Dropout \| P681H \| S477N \| T478K \| T95I \| Y144del_Composite |
| PV45494 | 12/14/21 | EPI_ISL_7908057 | BA.1 | D614G Detected. Omicron (B.1.1.529) Detected. | D614G \| H69_V70del_Dropout \| K417N \| N439K_Dropout \| P681H \| S477N \| T478K \| T95I \| Y144del_Composite |
| PV45437 | 12/14/21 | EPI_ISL_7908069 | BA.1 | D614G Detected. Omicron (B.1.1.529) Detected. | D614G \| H69_V70del_Dropout \| K417N \| N439K_Dropout \| P681H \| S477N \| T478K \| T95I \| Y144del_Composite |
| PV48991 | 12/19/21 | EPI_ISL_11178972 | AY.44 | D614G Detected. Delta (B.1.617.2) Detected. | D614G \| L452R \| P681R \| T19R \| T478K |
| PV47876 | 12/20/21 | EPI_ISL_11178970 | BA.1 | D614G Detected. | D614G \| H69_V70del_Dropout \| N439K_Dropout \| S477N \| T478K \| T95I |
| PV48993 | 12/21/21 | EPI_ISL_11178973 | AY.39.1 | D614G Detected. Delta (B.1.617.2) Detected. | D614G \| H69_V70del \| L452R \| P681R \| T19R \| T478K \| T95I |
| PV48978 | 12/25/21 | EPI_ISL_11178971 | AY.44 | D614G Detected. Delta (B.1.617.2) Detected. | D614G \| L452R \| P681R \| T19R \| T478K |
| PV49009 | 12/27/21 | EPI_ISL_11178974 | AY.109 | D614G Detected. Delta (B.1.617.2) Detected. | D614G \| H69_V70del \| L452R \| P681R \| T19R \| T478K \| T95I |
| PV49041 | 12/27/21 | EPI_ISL_11178975 | AY.3.3 | D614G Detected. Delta (B.1.617.2) Detected. | D614G \| H69_V70del \| L452R \| P681R \| T19R \| T478K |
| PV50567 | 1/3/22 | EPI_ISL_10831379 | BA.1 | D614G Detected. Omicron (B.1.1.529) Detected. | D614G \| H69_V70del_Dropout \| K417N \| N439K_Dropout \| P681H \| S477N \| T478K \| T95I \| Y144del_Composite |
| PV50565 | 1/3/22 | EPI_ISL_10831377 | BA.1 | D614G Detected. Omicron (B.1.1.529) Detected. | D614G \| H69_V70del_Dropout \| K417N \| N439K_Dropout \| P681H \| S477N \| T478K \| T95I \| Y144del_Composite |
| PV50564 | 1/3/22 | EPI_ISL_10831376 | BA.1.1 | D614G Detected. Omicron (B.1.1.529) Detected. | D614G \| H69_V70del_Dropout \| K417N \| N439K_Dropout \| P681H \| S477N \| T478K \| T95I \| Y144del_Composite |
| PV50568 | 1/3/22 | EPI_ISL_10831380 | BA.1.1 | D614G Detected. | D614G \| E484-K/Q_Dropout \| H69_V70del_Dropout \| N439K_Dropout \| S477N \| T478K \| T95I |
| PV50450 | 1/4/22 | EPI_ISL_10831369 | AY.43 | D614G Detected. Delta (B.1.617.2) Detected. | D614G \| L452R \| P681R \| T19R \| T478K |
| PV50577 | 1/4/22 | EPI_ISL_10831389 | BA.1.1 | D614G Detected. Omicron (B.1.1.529) Detected. | D614G \| H69_V70del_Dropout \| K417N \| N439K_Dropout \| P681H \| S477N \| T478K \| T95I \| Y144del_Composite |
| PV50569 | 1/4/22 | EPI_ISL_10831381 | BA.1.1 | D614G Detected. Omicron (B.1.1.529) Detected. | D614G \| H69_V70del_Dropout \| K417N \| N439K_Dropout \| P681H \| S477N \| T478K \| T95I \| Y144del_Composite |
| PV50571 | 1/4/22 | EPI_ISL_10831383 | BA.1.1 | D614G Detected. Omicron (B.1.1.529) Detected. | D614G \| H69_V70del_Dropout \| K417N \| N439K_Dropout \| P681H \| S477N \| T478K \| T95I \| Y144del_Composite |
| PV50451 | 1/4/22 | EPI_ISL_10831370 | BA.1 | D614G Detected. Omicron (B.1.1.529) Detected. | D614G \| H69_V70del_Dropout \| K417N \| N439K_Dropout \| P681H \| S477N \| T478K \| T95I \| Y144del_Composite |
| PV50489 | 1/4/22 | EPI_ISL_10831372 | BA.1.1 | D614G Detected. Omicron (B.1.1.529) Detected. | D614G \| H69_V70del_Dropout \| K417N \| N439K_Dropout \| P681H \| S477N \| T478K \| T95I \| Y144del_Composite |
| PV50615 | 1/4/22 | EPI_ISL_10831396 | BA.1.1 | D614G Detected. Omicron (B.1.1.529) Detected. | D614G \| H69_V70del_Dropout \| K417N \| N439K_Dropout \| P681H \| S477N \| T478K \| T95I \| Y144del_Composite |
| PV50488 | 1/4/22 | Mixed | BA.1.1 | D614G Detected. Omicron (B.1.1.529) Detected. | D614G \| H69_V70del_Dropout \| K417N \| N439K_Dropout \| P681H \| S477N \| T478K \| T95I \| Y144del_Composite |
| PV50574 | 1/4/22 | EPI_ISL_10831386 | BA.1.1 | D614G Detected. Omicron (B.1.1.529) Detected. | D614G \| H69_V70del_Dropout \| K417N \| N439K_Dropout \| P681H \| S477N \| T478K \| T95I \| Y144del_Composite |
| PV50570 | 1/4/22 | EPI_ISL_10831382 | BA.1 | D614G Detected. Omicron (B.1.1.529) Detected. | A701V \| D614G \| H69_V70del_Dropout \| K417N \| N439K_Dropout \| P681H \| S477N \| T478K \| T95I \| Y144del_Composite |
| PV50576 | 1/4/22 | EPI_ISL_10831388 | BA.1 | D614G Detected. Omicron (B.1.1.529) Detected. | A701V \| D614G \| H69_V70del_Dropout \| K417N \| N439K_Dropout \| P681H \| S477N \| T478K \| T95I \| Y144del_Composite |
| PV50501 | 1/4/22 | EPI_ISL_10831375 | BA.1.1 | D614G Detected. Omicron (B.1.1.529) Detected. | D614G \| H69_V70del_Dropout \| K417N \| N439K_Dropout \| P681H \| S477N \| T478K \| T95I \| Y144del_Composite |
| PV50575 | 1/4/22 | EPI_ISL_10831387 | BA.1 | D614G Detected. Omicron (B.1.1.529) Detected. | A701V \| D614G \| H69_V70del_Dropout \| K417N \| N439K_Dropout \| P681H \| S477N \| T478K \| T95I \| Y144del_Composite |
| PV50572 | 1/4/22 | EPI_ISL_10831384 | BA.1.1 | D614G Detected. Omicron (B.1.1.529) Detected. | D614G \| H69_V70del_Dropout \| K417N \| N439K_Dropout \| P681H \| S477N \| T478K \| T95I \| Y144del_Composite |
| PV50566 | 1/4/22 | EPI_ISL_10831378 | BA.1 | D614G Detected. Omicron (B.1.1.529) Detected. | D614G \| H69_V70del_Dropout \| K417N \| N439K_Dropout \| P681H \| S477N \| T478K \| T95I \| Y144del_Composite |
| PV50573 | 1/4/22 | EPI_ISL_10831385 | BA.1 | D614G Detected. Omicron (B.1.1.529) Detected. | D614G \| H69_V70del_Dropout \| K417N \| N439K_Dropout \| P681H \| S477N \| T478K \| T95I \| Y144del_Composite |
| PV50579 | 1/4/22 | EPI_ISL_10831391 | BA.1.1 | D614G Detected. Omicron (B.1.1.529) Detected. | D614G \| H69_V70del_Dropout \| K417N \| N439K_Dropout \| P681H \| S477N \| T478K \| T95I \| Y144del_Composite |
| PV50578 | 1/4/22 | EPI_ISL_10831390 | BA.1.1 | D614G Detected. Omicron (B.1.1.529) Detected. | D614G \| H69_V70del_Dropout \| K417N \| N439K_Dropout \| P681H \| S477N \| T478K \| T95I \| Y144del_Composite |
| PV50491 | 1/4/22 | EPI_ISL_10831374 | BA.1 | D614G Detected. Omicron (B.1.1.529) Detected. | D614G \| H69_V70del_Dropout \| K417N \| N439K_Dropout \| P681H \| S477N \| T478K \| T95I |
| PV50490 | 1/4/22 | EPI_ISL_10831373 | BA.1 | D614G Detected. Omicron (B.1.1.529) Detected. | D614G \| H69_V70del_Dropout \| K417N \| N439K_Dropout \| P681H \| S477N \| T478K \| T95I \| Y144del_Composite |
| PV50485 | 1/5/22 | EPI_ISL_10831371 | BA.1 | D614G Detected. Omicron (B.1.1.529) Detected. | D614G \| H69_V70del_Dropout \| K417N \| N439K_Dropout \| P681H \| S477N \| T478K \| T95I \| Y144del_Composite |
| PV50446 | 1/5/22 | EPI_ISL_10831365 | BA.1.1 | D614G Detected. Omicron (B.1.1.529) Detected. | D614G \| H69_V70del_Dropout \| K417N \| N439K_Dropout \| P681H \| S477N \| T478K \| T95I \| Y144del_Composite |
| PV50448 | 1/5/22 | EPI_ISL_10831367 | BA.1 | D614G Detected. Omicron (B.1.1.529) Detected. | D614G \| H69_V70del \| K417N \| N439K_Dropout \| P681H \| S477N \| T478K \| T95I \| Y144del_Composite |
| PV50449 | 1/5/22 | EPI_ISL_10831368 | BA.1 | D614G Detected. Omicron (B.1.1.529) Detected. | D614G \| H69_V70del_Dropout \| K417N \| N439K_Dropout \| P681H \| S477N \| T478K \| T95I \| Y144del_Composite |
| PV50444 | 1/5/22 | EPI_ISL_10831364 | BA.1 | D614G Detected. Omicron (B.1.1.529) Detected. | D614G \| H69_V70del_Dropout \| K417N \| N439K_Dropout \| P681H \| S477N \| T478K \| T95I \| Y144del_Composite |
| PV50429 | 1/10/22 | EPI_ISL_10831356 | BA.1 | D614G Detected. Omicron (B.1.1.529) Detected. | D614G \| H69_V70del_Dropout \| K417N \| N439K_Dropout \| P681H \| S477N \| T478K \| T95I \| Y144del_Composite |
| PV50436 | 1/10/22 | EPI_ISL_10831359 | BA.1.1 | D614G Detected. Omicron (B.1.1.529) Detected. | D614G \| H69_V70del_Dropout \| K417N \| N439K_Dropout \| P681H \| S477N \| T478K \| T95I \| Y144del_Composite |
| PV50430 | 1/10/22 | EPI_ISL_10831357 | BA.1.1 | D614G Detected. Omicron (B.1.1.529) Detected. | D614G \| H69_V70del_Dropout \| K417N \| N439K_Dropout \| P681H \| S477N \| T478K \| T95I \| Y144del_Composite |
| PV50608 | 1/10/22 | EPI_ISL_10831394 | BA.1.1 | D614G Detected. Omicron (B.1.1.529) Detected. | D614G \| H69_V70del_Dropout \| K417N \| N439K_Dropout \| P681H \| S477N \| T478K \| T95I \| Y144del_Composite |
| PV50609 | 1/11/22 | EPI_ISL_10831395 | BA.1 | D614G Detected. Omicron (B.1.1.529) Detected. | D614G \| H69_V70del_Dropout \| K417N \| N439K_Dropout \| P681H \| S477N \| T478K \| T95I \| Y144del_Composite |
| PV50433 | 1/11/22 | EPI_ISL_10831358 | BA.1 | D614G Detected. Omicron (B.1.1.529) Detected. | D614G \| K417N \| N439K_Dropout \| P681H \| S477N \| T478K \| T95I \| Y144del_Composite |
| PV50438 | 1/11/22 | EPI_ISL_10831360 | BA.1.1 | D614G Detected. Omicron (B.1.1.529) Detected. | D614G \| H69_V70del_Dropout \| K417N \| N439K_Dropout \| P681H \| S477N \| T478K \| T95I \| Y144del_Composite |
| PV50441 | 1/11/22 | EPI_ISL_10831362 | BA.1 | D614G Detected. Omicron (B.1.1.529) Detected. | D614G \| H69_V70del_Dropout \| K417N \| N439K_Dropout \| P681H \| S477N \| T478K \| T95I \| Y144del_Composite |
| PV50439 | 1/11/22 | EPI_ISL_10831361 | BA.1.1 | D614G Detected. Omicron (B.1.1.529) Detected. | D614G \| H69_V70del_Dropout \| K417N \| N439K_Dropout \| P681H \| S477N \| T478K \| T95I \| Y144del_Composite |
| PV50607 | 1/11/22 | EPI_ISL_10831393 | BA.1.1 | D614G Detected. Omicron (B.1.1.529) Detected. | D614G \| K417N \| N439K_Dropout \| P681H \| S477N \| T478K \| T95I |
| PV50605 | 1/11/22 | EPI_ISL_10831392 | BA.1.1 | D614G Detected. Omicron (B.1.1.529) Detected. | D614G \| H69_V70del_Dropout \| K417N \| N439K_Dropout \| P681H \| S477N \| T478K \| T95I \| Y144del_Composite |
| PV51975 | 1/24/22 | EPI_ISL_11178976 | BA.2 | D614G Detected. | D614G \| K417N \| N439K_Dropout \| N501Y \| P681H \| S477N \| T478K |
| PV55589 | 2/1/22 | EPI_ISL_11178988 | BA.2 | D614G Detected. | D614G \| K417N \| N439K_Dropout \| N501Y \| P681H \| S477N \| T478K |
| PV55575 | 2/1/22 | EPI_ISL_11178987 | BA.2 | D614G Detected. | D614G \| K417N \| N439K_Dropout \| N501Y \| P681H \| S477N \| T478K |
| PV56004 | 2/15/22 | EPI_ISL_11179017 | BA.2 | D614G Detected. | D614G \| N439K_Dropout \| N501Y \| P681H \| S477N \| T478K |
| PV56011 | 2/16/22 | EPI_ISL_11179020 | BA.2 | D614G Detected. | D614G \| K417N \| N439K_Dropout \| N501Y \| P681H \| S477N \| T478K |
| PV56107 | 2/22/22 | EPI_ISL_11179051 | BA.2 | D614G Detected. | D614G \| K417N \| N439K_Dropout \| N501Y \| P681H \| S477N \| T478K |
| PV56159 | 2/24/22 | EPI_ISL_11179065 | BA.2 | D614G Detected. | D614G \| K417N \| N439K_Dropout \| N501Y \| P681H \| S477N \| T478K |
| PV56250 | 2/25/22 | EPI_ISL_11179073 | BA.2 | D614G Detected. | D614G \| K417N \| N439K_Dropout \| N501Y \| P681H \| S477N \| T478K |
| PV56302 | 3/2/22 | EPI_ISL_11179077 | BA.2 | D614G Detected. | D614G \| N439K_Dropout \| N501Y \| P681H \| S477N \| T478K |

^a^ GISAID accession IDs are indicated for specimens with single variant consensus genomes; mixed assemblies are not deposited in GISAID.

^b^ Variant ID result from variant report output file.

^c^ SARS-CoV-2 amino acid polymorphisms separated by bars (“|”) per the variant report output file. Native amino acid calls are not listed if detected.

**Table S5. Predictive values for panel variant calls**

| **Variant** | **Number of Specimens** ^a^ | **PPV (95% CI)** | **NPV (95% CI)** | **P-value ^b^** |
| --- | --- | --- | --- | --- |
| Omicron (BA.1) | 79 | 1.000 (0.951-1.000) | 0.984 (0.964-0.993) | <0.0001 |
| Delta | 110 | 0.991 (0.949-1.000) | 0.986 (0.964-0.995) | <0.0001 |
| Alpha | 40 | 1.000 (0.912-1.000) | 1.000 (0.989-1.000) | <0.0001 |
| Beta | 4 | 1.000 (0.439-1.000) | 0.997 (0.986-1.000) | <0.0001 |
| Gamma | 14 | 0.933 (0.702-0.997) | 1.000 (0.990-1.000) | <0.0001 |
| Zeta | 1 | 0 (0.000-0.204) | 0.997 (0.985-1.000) | >0.999 |
| Eta | 7 | 1.000 (0.051-1.000) | 0.985 (0.967-0.993) | 0.0179 |
| Iota | 39 | 1.000 (0.908-1.000) | 0.997 (0.984-1.000) | <0.0001 |
| Epsilon | 19 | 1.000 (0.832-1.000) | 1.000 (0.990-1.000) | <0.0001 |
| B.1.258 | 1 | 1.000 (0.051-1.000) | 1.000 (0.990-1.000) | 0.0026 |
| D614G ^c^ | 27 | NA | NA | NA |
| Broad USA | 5 | 1.000 (0.566-1.000) | 1.000 (0.990-1.000) | <0.0001 |

^a^ Number of specimens confirmed by WGS as the indicated variant.

^b^ By Fisher’s exact test.

^c^ Agreement analyses were not performed as specimens that both (1) harbored the native D614 amino acid and (2) did not identify as any other variant on the panel were not recovered for testing. NA, not available.

**Table S6. Predictive values for panel target calls**

| **Target** | **Number of Specimens ^a, b^** | **PPV (95% CI)** | **NPV (95% CI)** | **P-value ^c^** |
| --- | --- | --- | --- | --- |
| L5F | 42 | 1.000 (0.912-1.000) | 0.994 (0.980-0.999) | <0.0001 |
| S13I | 19 | 1.000 (0.824-1.000) | 0.997 (0.985-1.000) | <0.0001 |
| L18F | 15 | 0.790 (0.567-0.915) | 1.000 (0.990-1.000) | <0.0001 |
| T19R | 110 | 0.991 (0.949-1.000) | 0.986 (0.964-0.995) | <0.0001 |
| H69_V70del | 51 | 0.922 (0.815-0.969) | 0.984 (0.960-0.994) | <0.0001 |
| D80A | 4 | 1.000 (0.439-1.000) | 0.997 (0.986-1.000) | <0.0001 |
| D80G | 0 | NA | NA | NA |
| T95I | 159 | 0.924 (0.874-0.955) | 0.996 (0.975-1.000) | <0.0001 |
| Y144del | 126 | 0.881 (0.813-0.927) | 0.943 (0.909-0.965) | <0.0001 |
| W152C | 19 | 1.000 (0.832-1.000) | 1.000 (0.990-1.000) | <0.0001 |
| D215G | 3 | 0 | 0.992 (0.978-0.998) | >0.9999 |
| L242_244del | 4 | 0.667 (0.119-0.983) | 0.995 (0.981-0.999) | 0.0002 |
| D253G | 39 | 0.927 (0.806-0.975) | 0.997 (0.984-1.000) | <0.0001 |
| K417N | 92 | 0.988 (0.937-0.999) | 0.977 (0.953-0.989) | <0.0001 |
| K417T | 14 | 0.824 (0.590-0.938) | 1.000 (0.990-1.000) | <0.0001 |
| N439K | 1 | 1.000 (0.051-1.000) | 1.000 (0.987-1.000) | 0.0034 |
| L452R | 124 | 0.961 (0.912-0.983) | 0.996 (0.979-1.000) | <0.0001 |
| Y453F | 0 | NA | NA | NA |
| S477N | 90 | 1.000 (0.959-1.000) | 1.000 (0.987-1.000) | <0.0001 |
| T478K | 194 | 0.980 (0.949-0.992) | 0.995 (0.971-1.000) | <0.0001 |
| E484Q | 0 | NA | NA | NA |
| E484K | 65 | 0.891 (0.791-0.946) | 0.974 (0.951-0.987) | <0.0001 |
| Q493K | 0 | NA | NA | NA |
| N501Y | 158 | 0.963 (0.898-0.990) | 0.744 (0.693-0.790) | <0.0001 |
| N501Y (exclude BA.1) | 79 | 0.963 (0.898-0.990) | 1.000 (0.984-1.000) | <0.0001 |
| N501T | 0 | NA | NA | NA |
| A570D | 40 | 1.000 (0.912-1.000) | 1.000 (0.989-1.000) | <0.0001 |
| D614G | 388 | 1.000 (0.990-1.000) | 0 (0.000-0.822) | >0.9999 |
| Q677H | 7 | 0.700 (0.397-0.892) | 1.000 (0.990-1.000) | <0.0001 |
| Q677P | 5 | 1.000 (0.566-1.000) | 1.000 (0.990-1.000) | <0.0001 |
| P681H | 141 | 0.986 (0.949-0.997) | 0.984 (0.960-0.994) | <0.0001 |
| P681R | 110 | 1.000 (0.964-1.000) | 0.979 (0.955-0.990) | <0.0001 |
| I692V | 0 | NA | NA | NA |
| A701V | 49 | 0.925 (0.821-0.970) | 1.000 (0.989-1.000) | <0.0001 |
| T716I | 40 | 1.000 (0.912-1.000) | 1.000 (0.989-1.000) | <0.0001 |
| S982A | 40 | 1.000 (0.910-1.000) | 0.997 (0.984-1.000) | <0.0001 |
| K1191N | 2 | 0.667 (0.119-0.983) | 1.000 (0.990-1.000) | <0.0001 |

^a^ Number of specimens that harbor the given target polymorphism by WGS.

^b^ Analyses were not performed if specimens with the given target polymorphism by WGS were not recovered for testing. NA, not available.

^c^ By Fisher’s exact test.
